# Impact of Australia’s 60-Day Dispensing Policy on Medicine Use: An Interrupted Time Series of the Stage 1 Roll-out

**DOI:** 10.64898/2026.09.09.26362679

**Authors:** Jack Janetzki, Nicole Pratt, Lachlan Dalli, Pilar Cataldo-Miranda, Sallie-Anne Pearson, Stella Talic, Lisa Kalisch Ellett

## Abstract

**Objective:** To evaluate the real-world impact of Stage 1 of Australia’s 60-day dispensing policy on medicine utilisation and contextualise changes in dispensing with medicine shortages.

**Methods:** We conducted an interrupted time series analysis of aggregated national monthly dispensing data from the Pharmaceutical Benefits Scheme and Repatriation Pharmaceutical Benefits Scheme for the 247 medicines included in Stage 1 of the 60-day dispensing policy. Data from January 2020 to October 2025 were analysed, with September 2023 defined as policy implementation. Linear regression assessed changes in monthly dispensing trends before and after implementation, accounting for autocorrelation and seasonal variation. A relative change of at least ±5% in the post-intervention dispensing slope compared with the pre-intervention slope was prespecified as a potentially clinically meaningful change. Therapeutic Goods Administration shortage reports were used to contextualise observed changes.

**Results:** Of 247 medicines, 131 (53%) had increasing and 116 (47%) had decreasing post-intervention dispensing trends; of which 86 (66%) and 71 (61%), were statistically significant, respectively. However, only 8 medicines (3%) demonstrated at least ±5% relative change in dispensing slope, six of which had reported shortages. Medicine-specific changes coincided with shortages, product deletions and therapeutic substitution, complicated attribution of changes to the 60-day dispensing policy.

**Conclusion:** Stage 1 of Australia’s 60-day dispensing policy was not associated with widespread changes in dispensing for most included medicines. Limited changes were plausibly influenced by concurrent shortages and other supply-related events. These findings provide reassurance that extended dispensing did not result in major increases in medicine utilisation at the population level, although continued monitoring is warranted as policy uptake expands.

**Short summary for non-experts:** Australia introduced 60-day dispensing in September 2023, allowing eligible patients to receive up to two months of some medicines at one time. We examined whether this changed overall usage of medicines across Australia.

We found that for almost all of the 247 medicines included in the first stage of the policy, there was no meaningful change in usage after 60-day dispensing was introduced. Only eight medicines showed a potentially meaningful change, and most of these changes occurred alongside medicine shortages or other supply problems. Overall, the findings suggest that 60- day dispensing did not lead to major increases in medicine usage, although ongoing monitoring is important as the policy expands.

## Introduction

The Pharmaceutical Benefits Scheme (PBS) is the primary mechanism for subsidising prescribed medicines in Australia, supporting affordable and equitable access to medicines.^1–3^ Recent cost-of-living pressures and concerns about medicine non-adherence have prompted reforms to improve medicine access.^3, 4^

A recent major reform was the introduction of 60-day dispensing in September 2023, allowing eligible patients with stable chronic conditions to receive up to 60 days of supply of specific PBS medicines for a single co-payment instead of the usual 30 days of supply.^4^ The co-payment refers to the cost a person would usually pay for a medicine each time it is dispensed at the pharmacy.^5^ In 2026, the co-payment is AU$25 per medicine for general PBS beneficiaries and AU$7.70 per medicine for concessional PBS beneficiaries who are eligible for social security payments.^4^ Stage 1 of the 60-day dispensing policy commenced on 1 September 2023 and included 247 medicines, primarily targeting medicines to treat chronic conditions such as cardiovascular disease and osteoporosis.^4^ Stages 2 and 3 of the policy commenced in March 2024 and September 2024 respectively, with additional medicines added on a rolling basis thereafter.^3^

A pre-implementation impact analysis of the policy by the Australian Government mentioned that less frequent dispensing of medicines would not affect overall medicine use as total quantities of medicines provided remain unchanged.^3^ However, whether this assumption was reflected in real-world practice is unclear.

Several Australian studies on other medicine policies related to opioids^2, 6, 7^, alprazolam^8^ and quetiapine^9^ have demonstrated that dispensing restrictions can significantly change medicine use patterns and may lead to unintended consequences. For this reason, evaluation is needed to monitor the intended and unintended changes with the 60-day dispensing policy, particularly given the assertion of the pre-implementation impact analysis. The aim of this study was to test the hypothesis that 60-day dispensing does not impact overall dispensing of medicines included in the policy Stage 1 and to contextualise observed changes in relation to reported medicine shortages.

## Methods

### Data sources

This study used aggregated monthly medicine dispensing data from the Pharmaceutical Benefits Scheme (PBS) and Repatriation Pharmaceutical Benefits Scheme (RPBS) Section 85 and Section 100 Date of Supply dataset.^10^ This dataset reports the number of prescriptions dispensed per month by PBS item code, corresponding to specific medicine strength, formulation, and pack size at a nationwide aggregate level. PBS item codes were linked to the PBS item drug map to identify corresponding medicine names.^10^

### Medicines of interest

Medicines in this study included those listed in Stage 1 of the 60-day dispensing policy. For each medicine, we extracted monthly dispensing counts for both the newly introduced 60-day item codes and the corresponding 30-day item codes. In the absence of individual-level data, we assumed people consumed one tablet per day of their medicine if it was dispensed in a quantity of 30 units. We assumed each 60-day supply represented two months of therapy, while each 30-day supply represented one month. This assumption has been used in prior analyses including those of the 60-day dispensing program.^11, 12^

### Study period and intervention

The study period was from January 2020 to October 2025. We defined September 2023 as the intervention point, corresponding to implementation of Stage 1 of the 60-day dispensing policy.

### Data analysis

We used an interrupted time series (ITS) design with linear regression to evaluate changes in dispensing associated with policy implementation. We performed analyses using SAS OnDemand for Academics. The primary outcome was the change in trend (slope change) of monthly dispensing rates following September 2023.

Using the dispensings per month, we fitted linear regression models to estimate pre-intervention trends, immediate level changes, and post-intervention trend changes. We addressed autocorrelation using the SAS AUTOREG procedure, with Durbin–Watson estimates and four quarterly dummy variables incorporated to account for seasonal variation. We assessed adequate adjustment for seasonality based on a Durbin–Watson statistic approaching two and the total R-square of the model estimates. The pre-intervention time period was January 2020 to August 2023 (44 months), the intervention month was September 2023) and the post-intervention time period was October 2023 to October 2025 (25 months). We treated September 2023 as a transition month and removed it from the ITS models. We did not assess the change in level (immediate effect) of monthly dispensing rates in September 2023 as prior analyses had shown that there was low initial uptake of policy.^13, 14^ We assessed the statistical significance of the post-intervention trend by comparing its slope with that of the pre-intervention trend, with p≤0.05 indicating a statistically significant difference.

To assess the magnitude of changes in medicine utilisation following implementation of Stage 1 of the 60-day dispensing policy, a relatively change of at least ±5% in monthly dispensing compared to baseline was prespecified as a threshold for a potentially clinically meaningful change in utilisation.

Three cardiovascular medicines were randomly selected from the pool of all Stage 1 medicines to visualise time series and ITS results: irbesartan 300 mg tablets, perindopril arginine 5 mg tablets (both antihypertensive agents used in management of hypertension), and a fixed-dose combination of ezetimibe 10 mg with atorvastatin 40 mg (a lipid-lowering therapy used for dyslipidaemia and cardiovascular risk reduction).

Prior to implementation of the policy, there were concerns that medication shortages could be associated with the policy or could be worsened with 60-day dispensing. To contextualise observed dispensing patterns, we obtained medicine shortage data from the Therapeutic Goods Administration Medicine Shortages Database, including both active and archived reports.^15^ We extracted the impact of reported shortages, as described in the TGA database, to aid in the contextualisation of the impact of the shortages on dispensing patterns.

### Ethics approval

Ethics approval was not required as the study used publicly available, aggregated, and de-identified data.

## Results

We included 247 medicines in our analysis. Prior to introduction of the policy change in September 2023, the largest increasing rate of dispensings per month was 2852 and the largest decreasing rate was −578 per month (IQR 149, 25^th^-75^th^ −15 to 135).

After policy introduction, the maximum monthly increase in dispensings was 4457 and the largest decrease was −4973 per month (IQR 69, 25^th^-75^th^ −35 to 34). Each month after September 2023, there was an increase in dispensings for 131 medicines (53%) of which, 86 (66%) were statistically significant increases. Each month after September 2023, there was a reduction in dispensing for 116 medicines (47%) of which, 71 (61%) were statistically significant.

A change in post-intervention dispensing slope of at least ±5% compared to baseline was observed for 8 of 247 medicines (3%); mesalazine 1.2g modified release 120 tablets, atenolol 50mg/mL oral liquid 300mL, mesalazine 3g modified release granules 30 sachets, ezetimibe 10mg 30 tablets, ezetimibe 10mg with atorvastatin 80mg 30 tablets, rivaroxaban 2.5mg 60 tablets, clopidogrel 75mg 28 tablets and mesalazine 1.6g enteric coated tablets 60 tablets. Of note, however, three of these medicines were added to the PBS during the study period impacting the interpretability of these results because of insufficient time prior to intervention. Mesalazine 1.2g modified release tablet 120 tablets were added to PBS in April 2023. Rivaroxaban 2.5mg 60 tablets was added in December 2020, and mesalazine 1.6g enteric coated tablets 60 tablets was added in May 2021.

Of these eight medicines/formulations however, mesalazine 1.2g modified release 120 tablets and mesalazine 3mg modified release granules 30 sachets had no reported shortages in the study period. Atenolol 50mg/mL oral liquid 300mL had two shortages; April 2020 to July 2020 and March 2021 to February 2022. Ezetimibe 10mg had 22 shortages over the study period. Ezetimibe 10mg with atorvastatin 80mg had 8 shortages. Rivaroxaban 2.5mg 60 tablets had one shortage from March 2025 to June 2025. Clopidogrel 75mg had 35 shortages over the study period and mesalazine 1.6g enteric coated tablets 60 tablets had 5 shortages.

Results for the three cardiovascular medicines that were selected to visualise time series are shown in Figures 1-3 and a summary of corresponding ITS results for these medicines is provided in Table 2. ITS model fit, measured by R^2^ was acceptable for all three of these medicines (Table 2). For irbesartan 300mg tablets (Figure 1) the monthly change in dispensing rates after the 60-day dispensing implementation was statistically significant, however, the change was not considered to be clinically meaningful given there was a reduction of 142 dispensings per month, representing 0.11% of baseline dispensings. While there were several shortages of irbesartan 300mg tablet products during the study period and two products deleted from the market (Appendix Table 1), the TGA classified all these shortages as low impact.^15^

**Figure 1.**
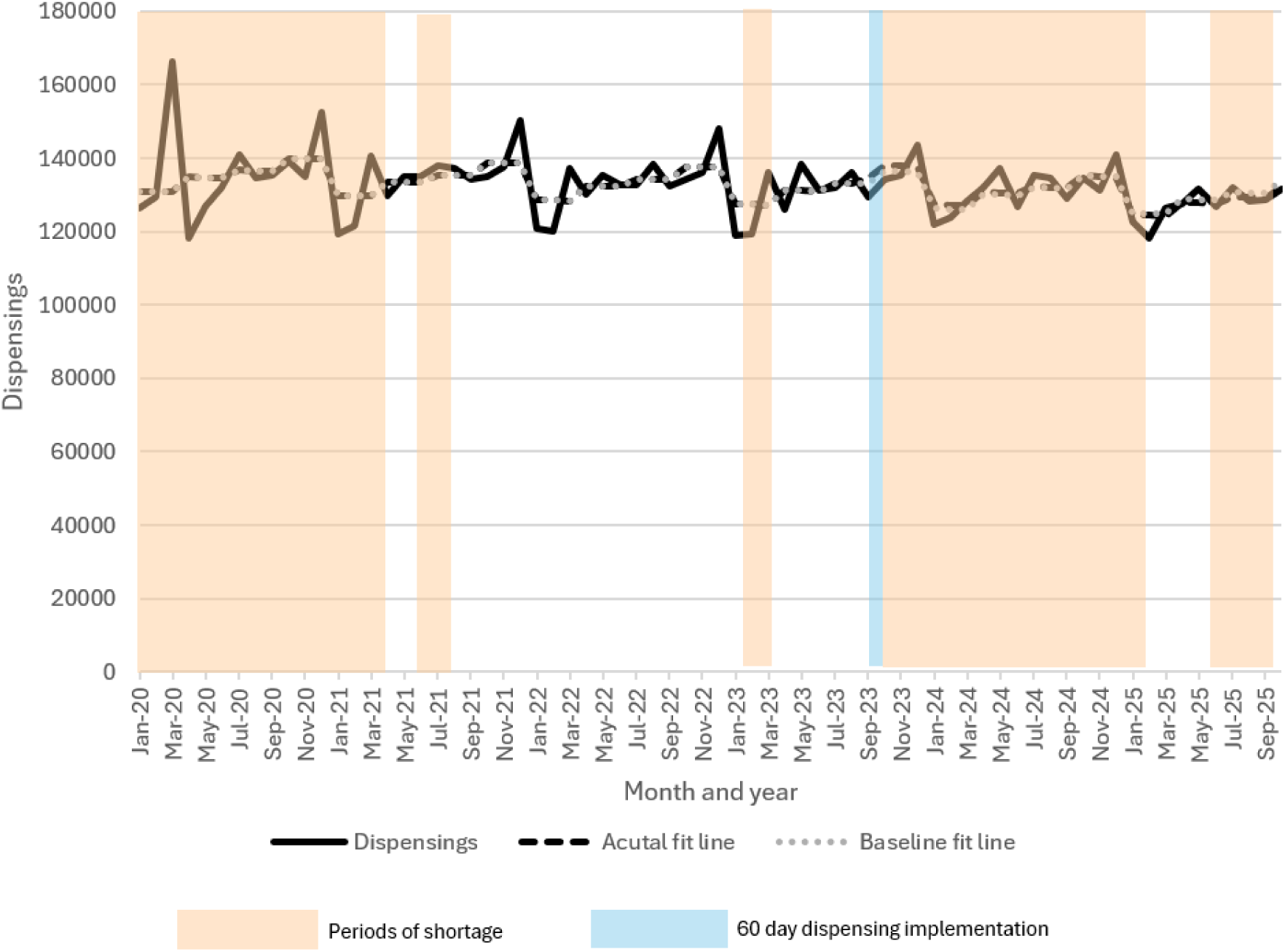
Irbesartan 300 mg tablet dispensings per month from January 2020 to October 2025. Blue line indicates implementation of 60-day dispensing in September 2023

For ezetimibe 10mg with atorvastatin 40mg tablets (Figure 2, Table 2), there was a statistically significant reduction of 947 dispensings per month in the months after policy implementation, however this was also considered to not be clinically meaningful as it represented 4.3% of baseline dispensings. Four shortages of ezetimibe 10mg with atorvastatin 40mg tablet products occurred throughout the study period (Appendix Table 2). The shortage from June 2020 to February 2021 was rated as medium impact by the TGA whereas the one in January 2024 was low impact. The first year long shortage from April 2024 to April 2025 was rated as medium impact whereas the shortage from July 2024 to July 2025 was rated as low impact.

**Figure 2.**
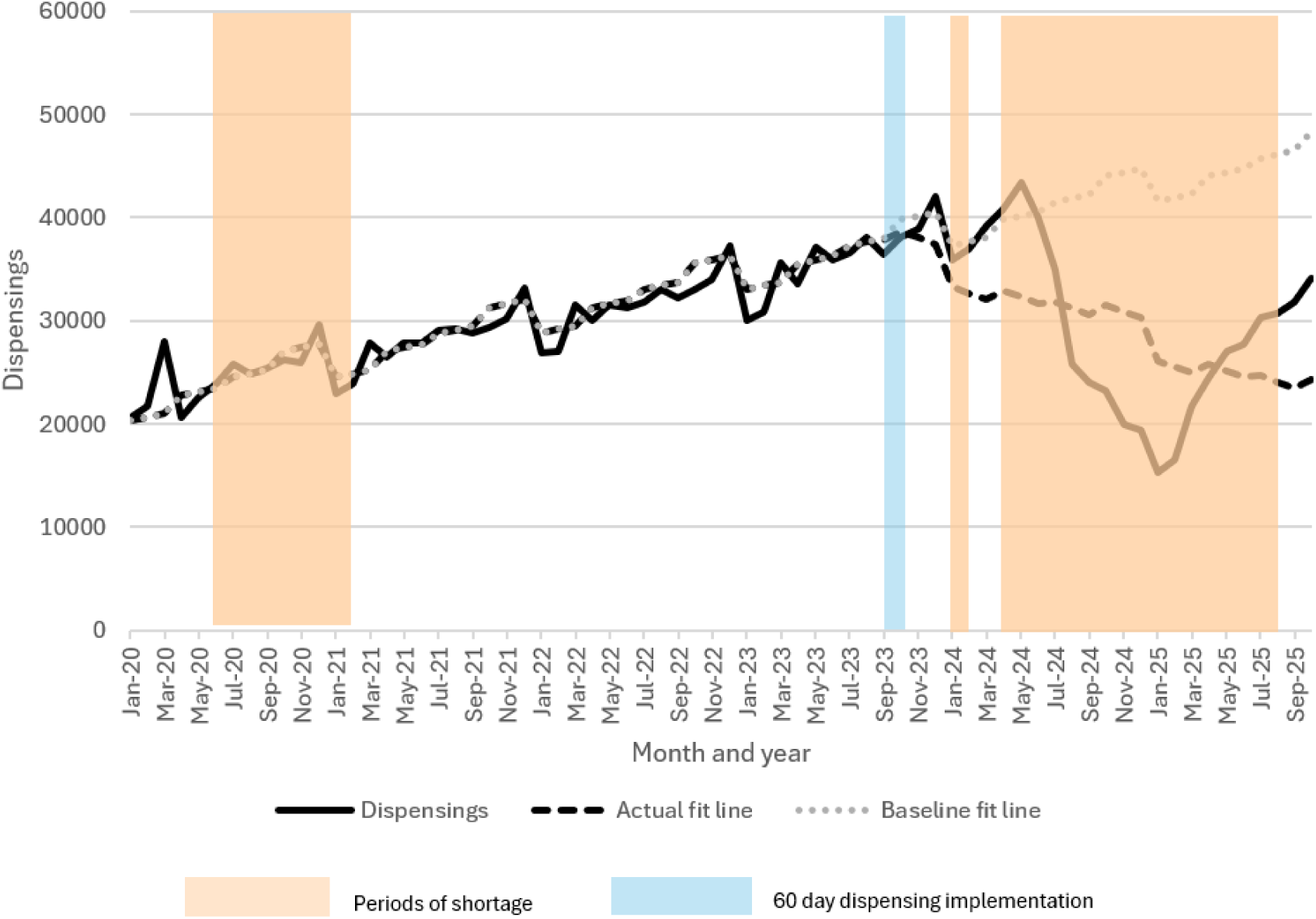
Ezetimibe 10 mg with atorvastatin 40 mg tablet dispensings per month from January 2020 to October 2025. Blue line indicates implementation of 60-day dispensing in September 2023. Orange bars indicate shortage periods of this combination medicine.

For perindopril arginine 5mg tablets (Figure 3, Table 2), the change in monthly dispensings was statistically significant following 60-day dispensing implementation. This change, however, was not considered clinically meaningful given that it represented 2.5% of baseline dispensings. Several shortages of perindopril erbumine 4mg or perindopril arginine 5mg products occurred throughout the study period. Figure 3 is overlayed with shortages of both as they are considered bioequivalent and interchangeable on the PBS. The immediate change in perindopril 5mg dispensings at policy implementation aligned with a shortage of perindopril at the same time.

**Figure 3.**
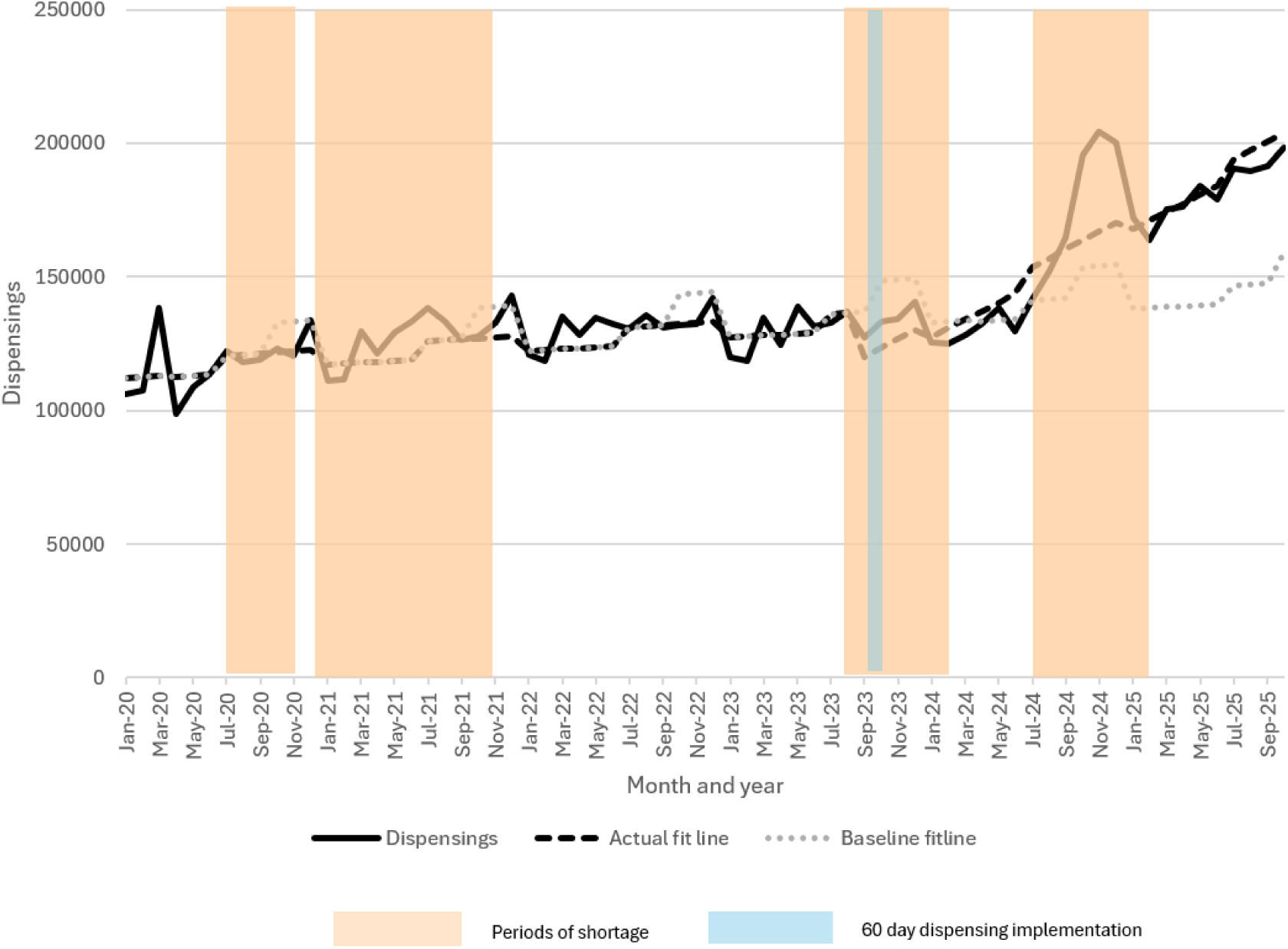
Perindopril arginine 5 mg dispensings per month from January 2020 to October 2025. Blue line indicates implementation of 60-day dispensing in September 2023. Orange bars indicate shortage periods of perindopril erbumine 4 mg or perindopril arginine 5 mg which are considered interchangeable and equivalent on the Pharmaceutical Benefits Scheme.

**Table 2:** Interrupted time series statistics for irbesartan 300mg tablet, ezetimibe 10mg with atorvastatin 40mg tablet and perindopril 5mg tablet.

| **Irbesartan 300mg tablet** |  |  |  |  |
| --- | --- | --- | --- | --- |
| **Term** | **Total R<sup>2</sup>** | **Dispensings per month** | **SE** | **P-value** |
| Dispensings/month in Jan 2020 | 0.6837 | 135207 | 1306 | <.0001 |
| Monthly change pre-September 2023 |  | -99.2 | 19.6 | <.0001 |
| Change in September 2023 |  | 1980 | 1015 | 0.057 |
| Monthly change post- September 2023 |  | -142.2 | 57.0 | 0.0160 |
| **Ezetimibe 10mg with atorvastatin 40mg tablet** |  |  |  |  |
| **Term** | **Total R<sup>2</sup>** | **Dispensings per month** | **SE** | **P-value** |
| Dispensings/month in Jan 2020 | 0.8687 | 22055 | 1635 | <0.0001 |
| Monthly change pre-September 2023 |  | 353.9 | 54.3 | <0.0001 |
| Change in September 2023 |  | 701.3 | 2350 | 0.77 |
| Monthly change post- September 2023 |  | -947.9 | 144.7 | <0.0001 |
| **Perindopril 5mg tablet** |  |  |  |  |
| **Term** | **Total R<sup>2</sup>** | **Dispensings per month** | **SE** | **P-value** |
| Dispensings /month in Jan 2020 | 0.9232 | 117558 | 3783 | <0.0001 |
| Monthly change pre-September 2023 |  | 432.5 | 108.8 | 0.0002 |
| Change in September 2023 |  | -19797 | 4830 | 0.0001 |
| Monthly change post- September 2023 |  | 2920 | 271.8 | <0.0001 |

A summary of the interrupted time series regression outputs for each medicine can be found in Appendix Table 5.

## Discussion

This study evaluated the real-world impact of Stage 1 of Australia’s 60-day dispensing policy on medicine use. Consistent with policy expectations, for most medicines there was no clinically meaningful change in monthly dispensings after policy implementation. For eight medicines there were clinically meaningful differences of at least ±5% change from baseline, however these were plausibly explained by concomitant events such as medicine shortages. Additionally, three of the eight medicines were added to the PBS during the study period, potentially resulting in a false positive association with significant change in trend post intervention as there may not have been sufficient pre-intervention time observed.

### Policy implementation and medicine shortages

A key concern prior to implementation of 60-day dispensing was the potential for medicine shortages to affect access to medicines included in the program. Our findings suggest that Stage 1 implementation of 60-day dispensing did not lead to major changes in dispensing trends at the population level. This may alleviate concerns raised by stakeholders regarding unintended increases in medicine use or stockpiling. However, these findings should also be interpreted in the context of the gradual and limited uptake of 60-day dispensing during the study period as the policy has not been universally adopted across eligible populations, prescribers, or medicines.^14^ Consequently, the observed population-level effects may reflect both the true impact of the policy and the extent of its implementation and uptake.

The medicine specific analyses for ezetimibe 10mg with atorvastatin 40mg and perindopril arginine 5mg illustrate that supply disruptions can influence dispensing patterns. The case study of perindopril arginine 5mg demonstrates how shortages, brand deletions, and therapeutic substitution can produce abrupt changes in dispensing independent of policy implementation. In particular, the increase in perindopril 5mg dispensings after September 2023 aligned with shortages and deletion of some brands of perindopril erbumine 4mg tablets from the PBS and likely switching to perindopril arginine. The increase in utilisation observed towards the end of 2024 and early 2025 is likely attributable to deletion of three perindopril erbumine 4mg products and sustained shortages of perindopril erbumine products (Appendix Table 3 and Appendix Table 4).

Irbesartan dispensings remained relatively stable despite multiple shortages classified as low impact, suggesting that availability of alternative products and the severity of supply disruptions play a critical role in shaping dispensing patterns. These findings emphasise the challenges of causal attribution in interrupted time series analyses undertaken during periods of substantial market disruption. Concurrent events including medicine shortages, brand discontinuations, therapeutic substitution, and changes in wholesaler or manufacturer supply can independently alter dispensing patterns and may confound attribution of observed changes solely to policy implementation. Consequently, estimates of the impact of 60-day dispensing should be interpreted within the broader context of these contemporaneous disruptions to medicine access and supply.

International evidence on extended dispensing suggests that there is improved adherence to medicines, minimal impact of policy on overall use of medicines and potential short term supply pressures when such policies are implemented. One systematic review including nine studies, rated as having moderate quality evidence, found that 2-4 month prescriptions improved adherence compared to one-month prescriptions.^16^ A study conducted in Thailand also found that increasing prescription lengths from one month to three months increased medication possession and was associated with improved disease markers in diabetes and dyslipidemia.^17^ There is however limited evidence on how extended dispensing intervals impact medicine waste and whether they contribute to are influenced by medicine shortages. There is some weak evidence to suggest that extended dispensing intervals may slightly increase unused medicine and wastage, which is important to highlight in the context of medicine shortages.^13, 16, 18, 19^ Unused medicine could otherwise be allocated to people impacted by shortages.^20^ Extended prescription durations have, however, been associated with cost savings and greater quality-adjusted life-years for patients.^18^

### Strengths and limitations

The use of aggregate national dispensing in this study allowed for population-level evaluation of policy implementation but prevented any person-level inferences.

Concurrent events such as medicine shortages and PBS listing changes (additions and deletions) may confound policy effects estimated from our ITS model that cannot be fully disentangled. Additionally, the pre-intervention period coincided with the COVID-19 pandemic which may have influenced dispensing patterns and the underlying pre-intervention period. It is important to note however that most medicines in Stage 1 of the 60-day dispensing policy are medicines for chronic conditions and people were able to visit pharmacies throughout the pandemic to access medicines. There were also several co-payment changes and revisions made to safety net thresholds throughout the study period.^3^ It is not possible to disaggregate the impacts of the policy change versus other interventions that happened concurrently. There is no established threshold for determining a clinically meaningful change in medicine dispensing, and the 5% threshold used in this study should therefore not be interpreted as a validated clinical threshold. Rather, it was selected a prior to identify changes of sufficient magnitude to be potentially import from a medicine utilisation and policy perspective. The findings of this study do, however, provide reassurance that Stage 1 of 60-day dispensing did not lead to great inappropriate increases in medicine use at a population level and supports the continued rollout of extended dispensing models. Ongoing surveillance remains important, particularly as additional medicines are incorporated into the policy and as medicine shortages continue to affect access to medicines in Australia.

## Conclusion

Although dispensing rates for medicines changed over time, for most medicines there was no clinically meaningful change in monthly dispensing following introduction of 60-day dispensing. These findings may alleviate concerns about unintended increases in medicine utilisation however the limited change in absolute dispensing rates is plausibly attributed to slow policy uptake, supply disruptions and other simultaneous changes to PBS listing of medicines. Continued monitoring of medicine utilisation in the context of extended dispensing intervals and shortages is warranted especially as the policy expands to additional medicines and while medicine shortages persist.

## Ethics and participant consent

This study used publicly available aggregate dispensing data therefore ethics approval was not required.

## Data availability

Data for this study are freely and publicly available from: https://www.pbs.gov.au/info/statistics/dos-and-dop/dos-and-dop

## Author contributions

Jack Janetzki: Conceptualisation, methodology, software, formal analysis, investigation, resources, data curation, writing – original draft, visualization. Project management

Nicole Pratt: Conceptualisation, methodology, funding acquisition, supervision, writing – review and editing, formal analysis

Lachlan Dalli: supervision, writing – review and editing

Pilar Cataldo-Miranda: supervision, writing – review and editing

Sallie-Anne Pearson: Funding acquisition, supervision, writing – review and editing

Stella Talic: supervision, writing – review and editing

Lisa Kalisch Ellett: Conceptualisation, methodology, supervision, writing – review and editing, formal analysis

## Acknowledgements

The authors acknowledge the discussions held with the NHMRC Medicines Intelligence Centre of Research Excellence 60-day dispensing program members and stakeholders in preparation of this manuscript.

## Appendix

**Appendix Table 1:**
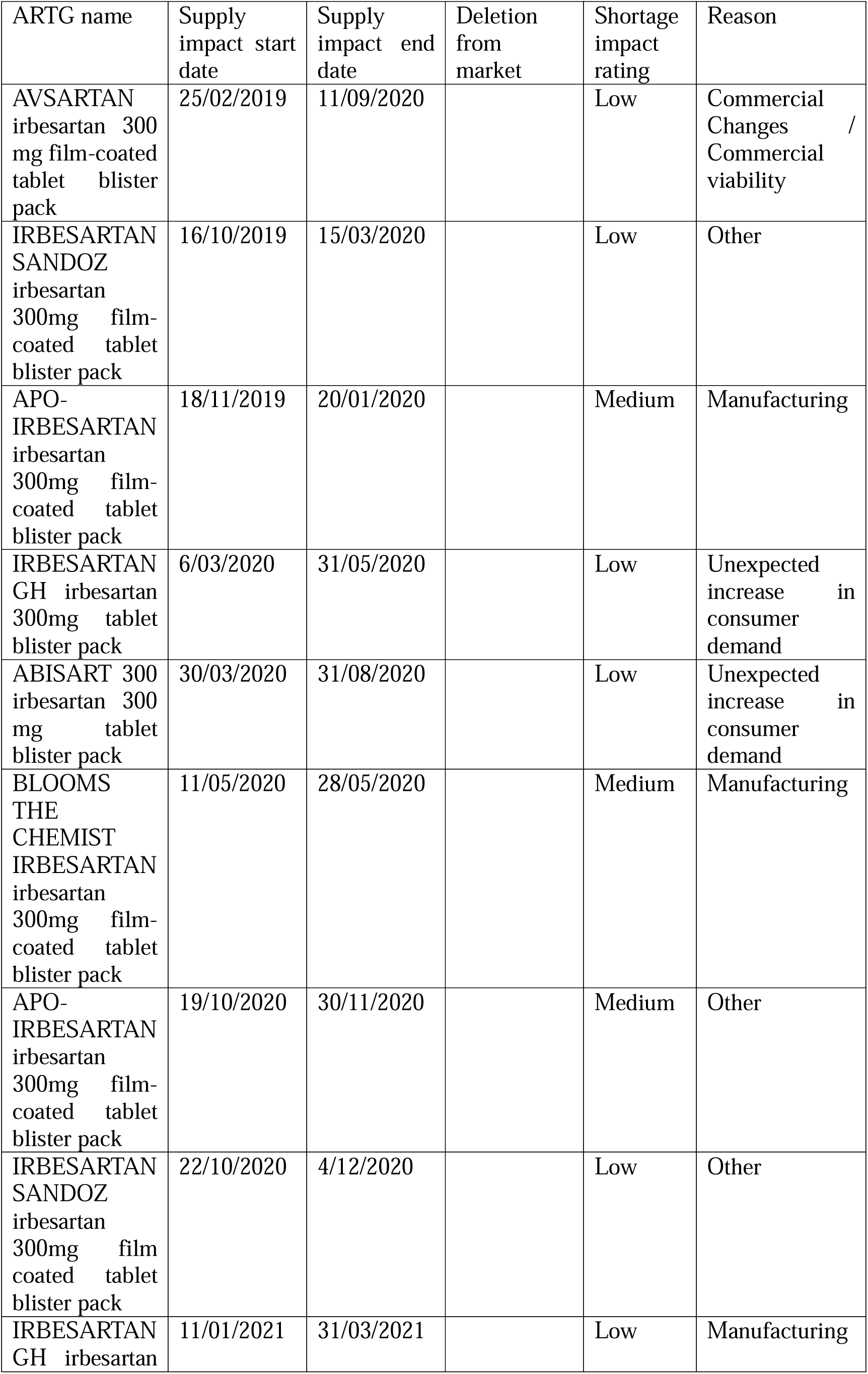
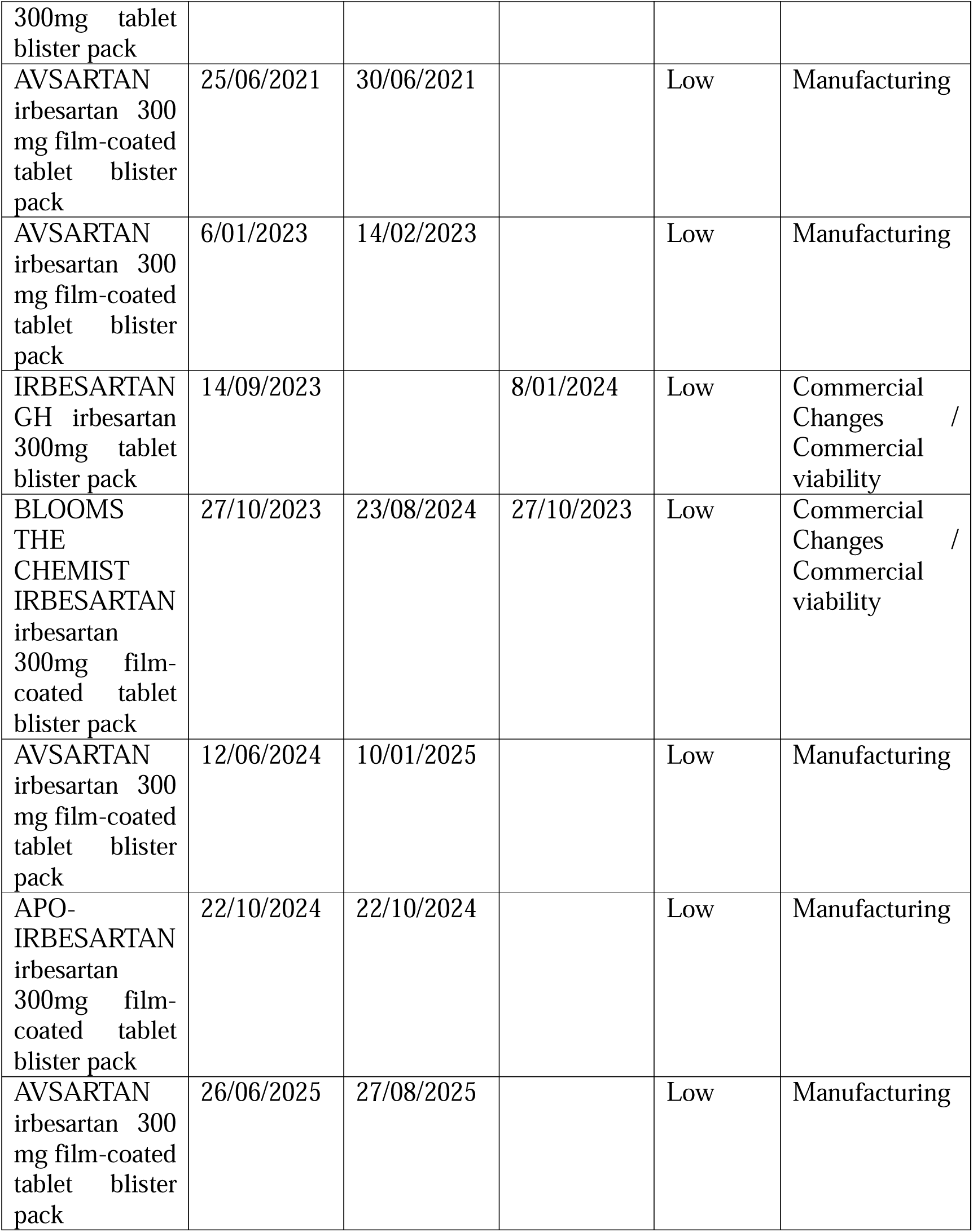
Irbesartan shortages and deletions:

**Appendix Table 2:**
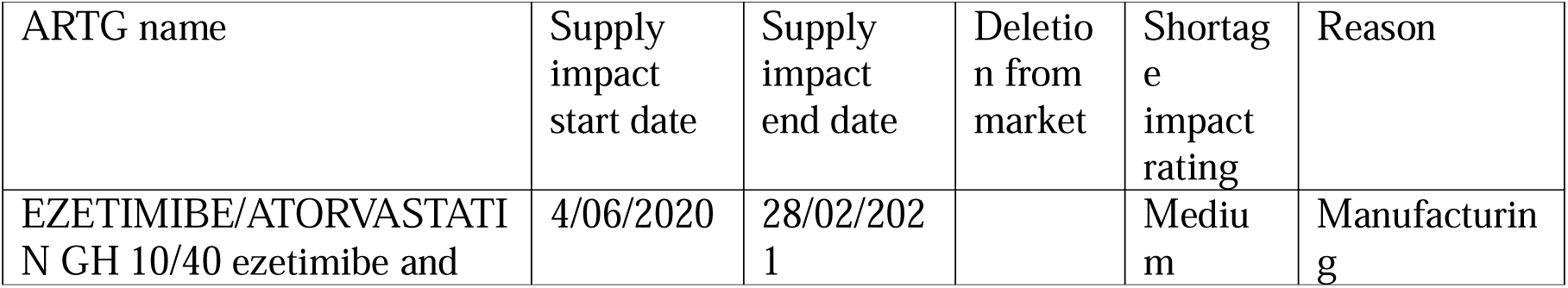
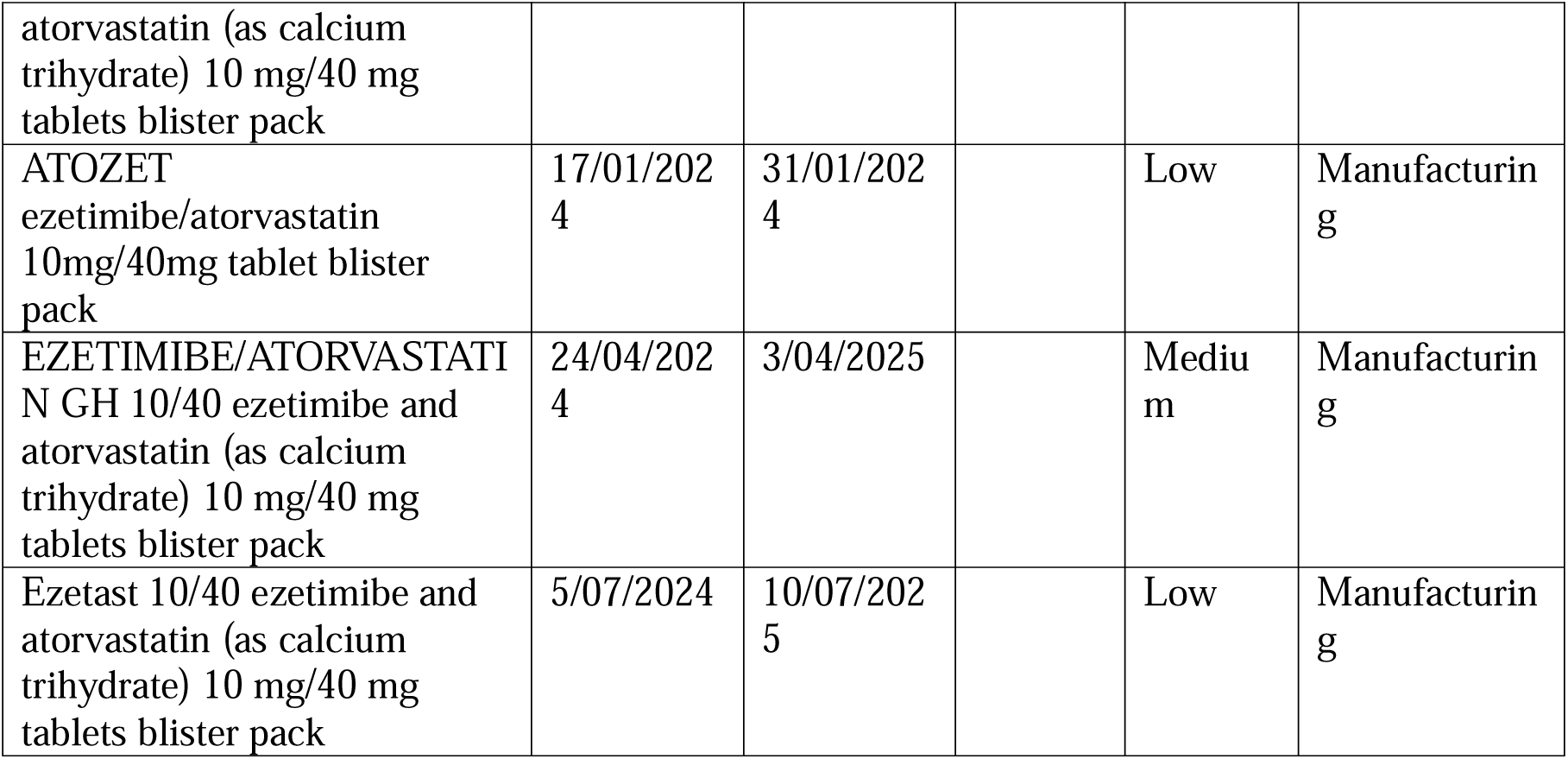
ezetimibe 10mg with atorvastatin 40mg tablet shortages.

**Appendix Table 3:**
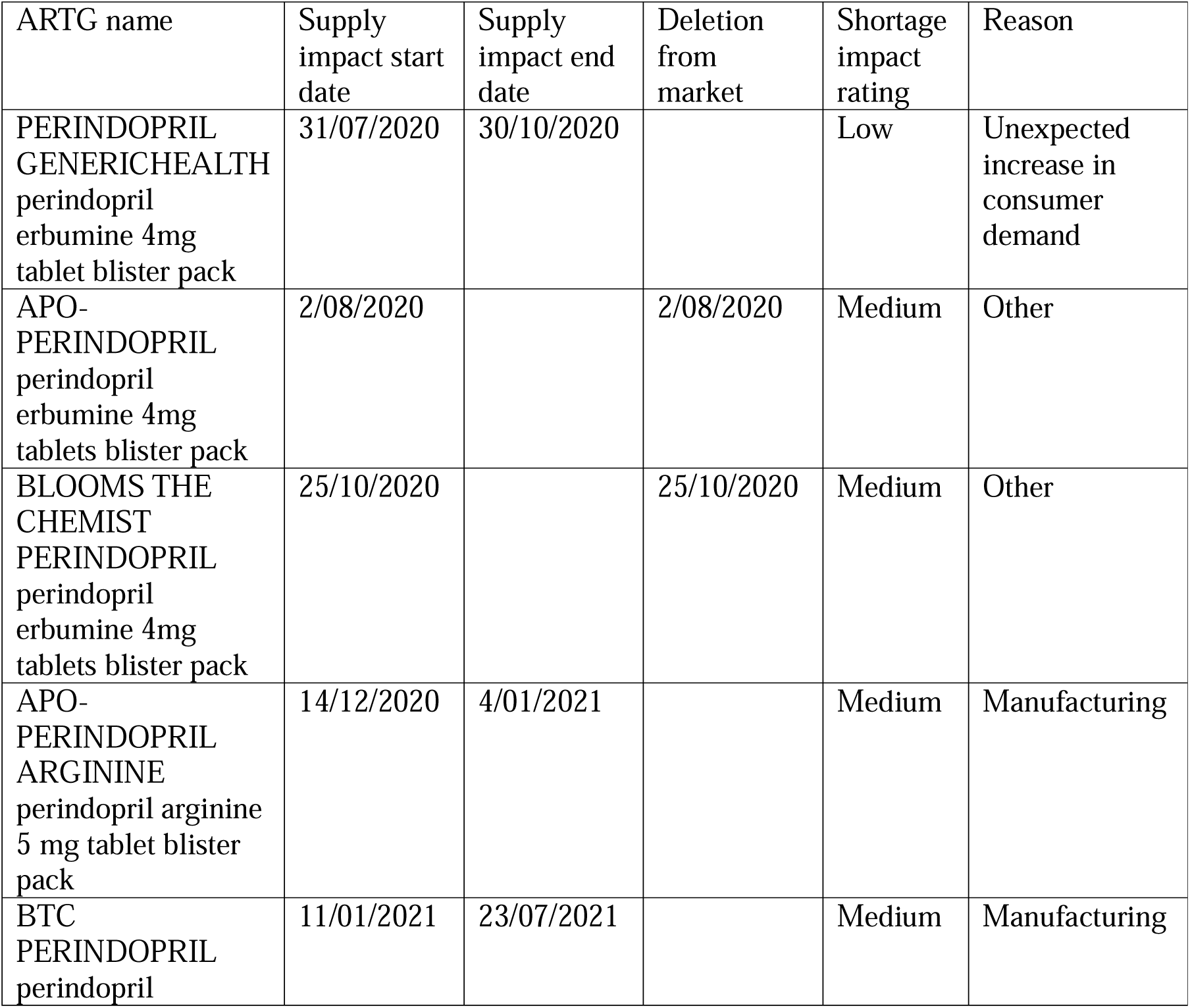
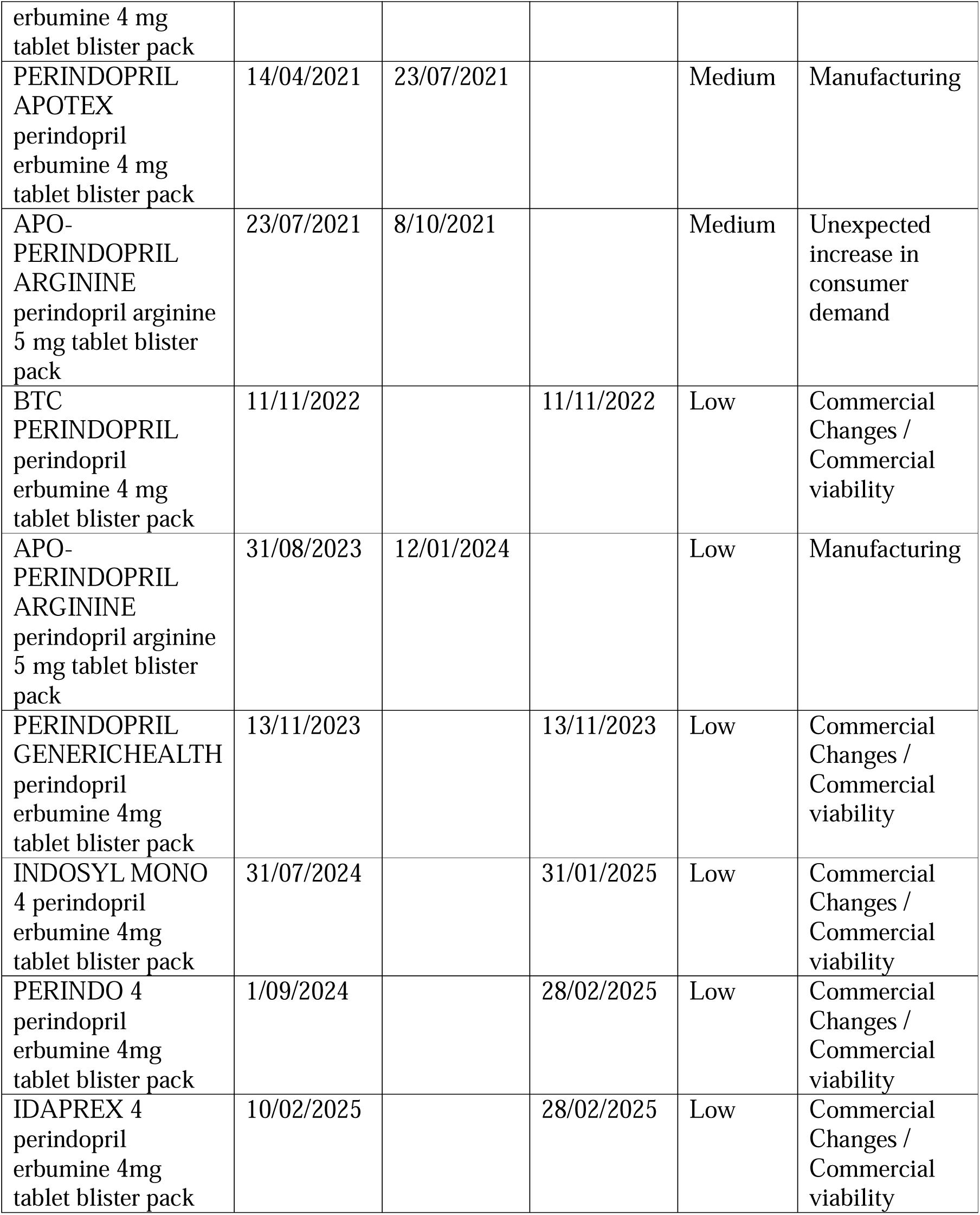
Perindopril shortages or deletions (includes both perindopril erbumine 4 mg and perindopril arginine 5 mg forms as they are considered interchangeable and bioequivalent on the PBS)

**Appendix Table 4.**
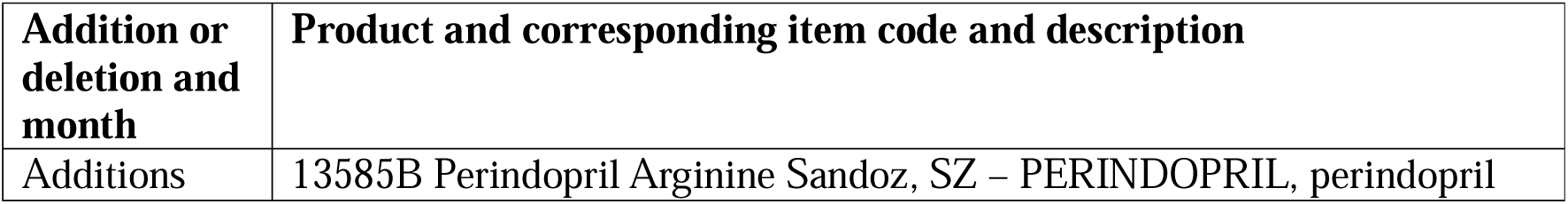
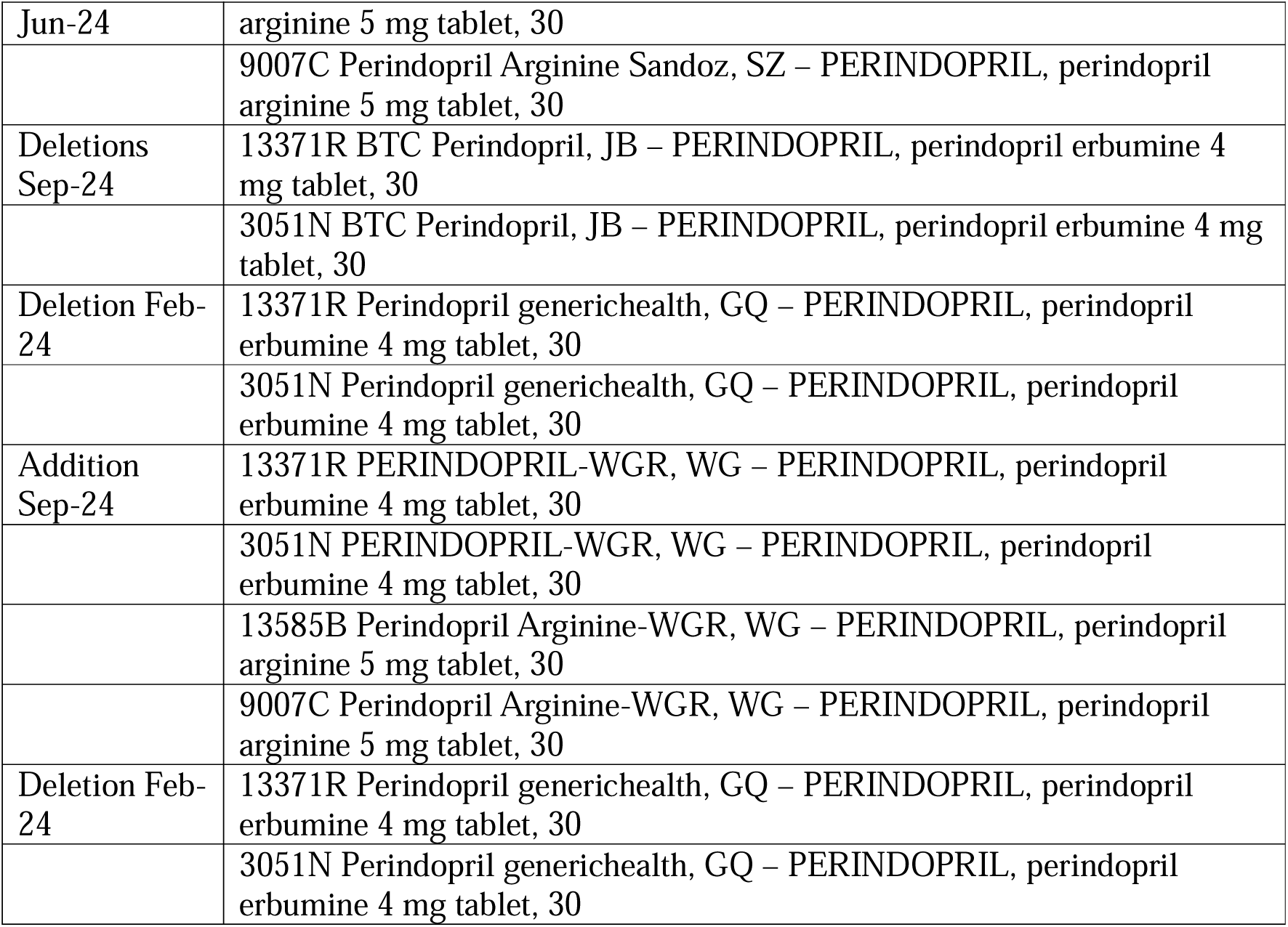
Additions or deletions of perindopril arginine 5mg and perindopril erbumine 4mg products from the PBS during the study period.

**Appendix Table 5.**
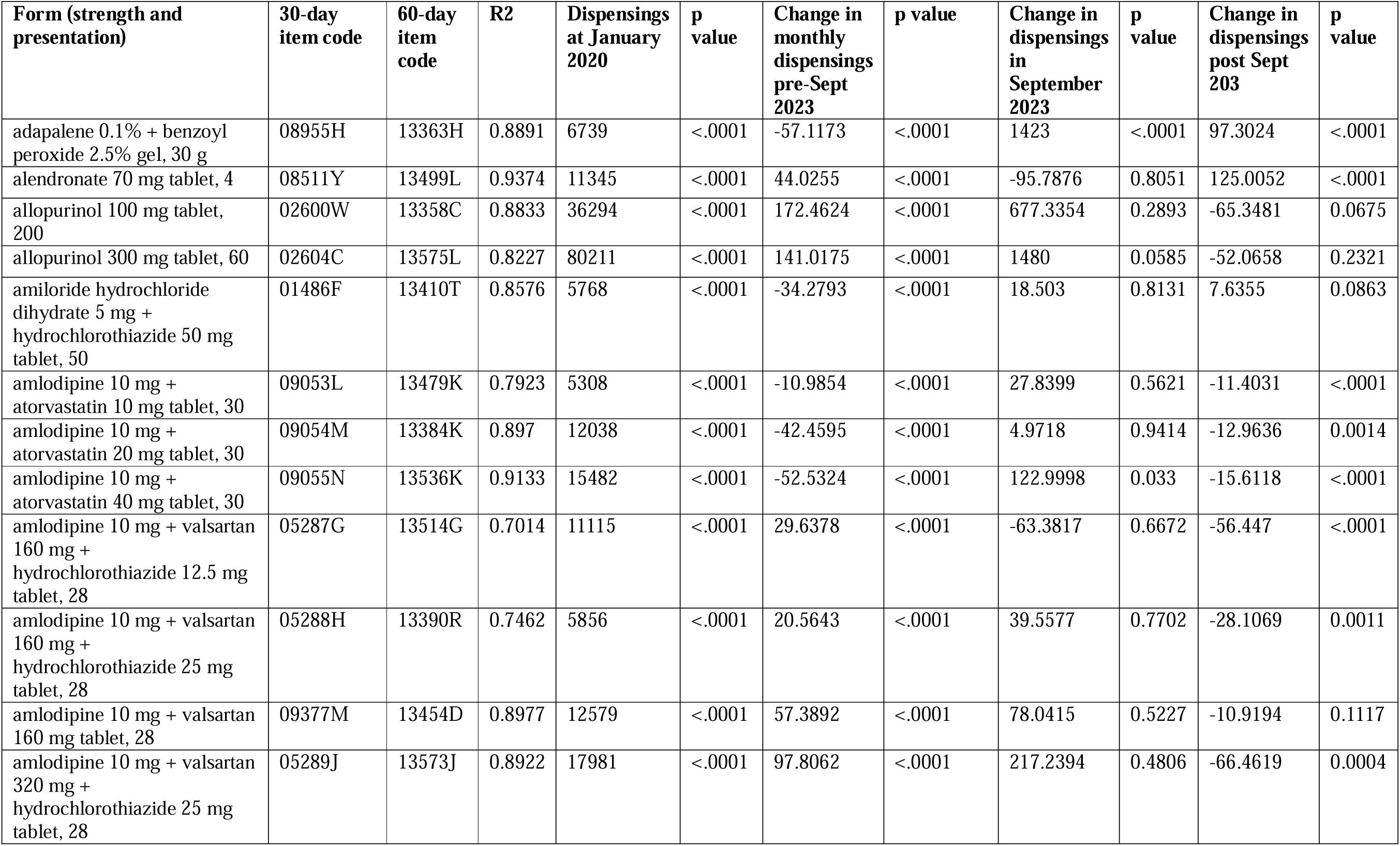
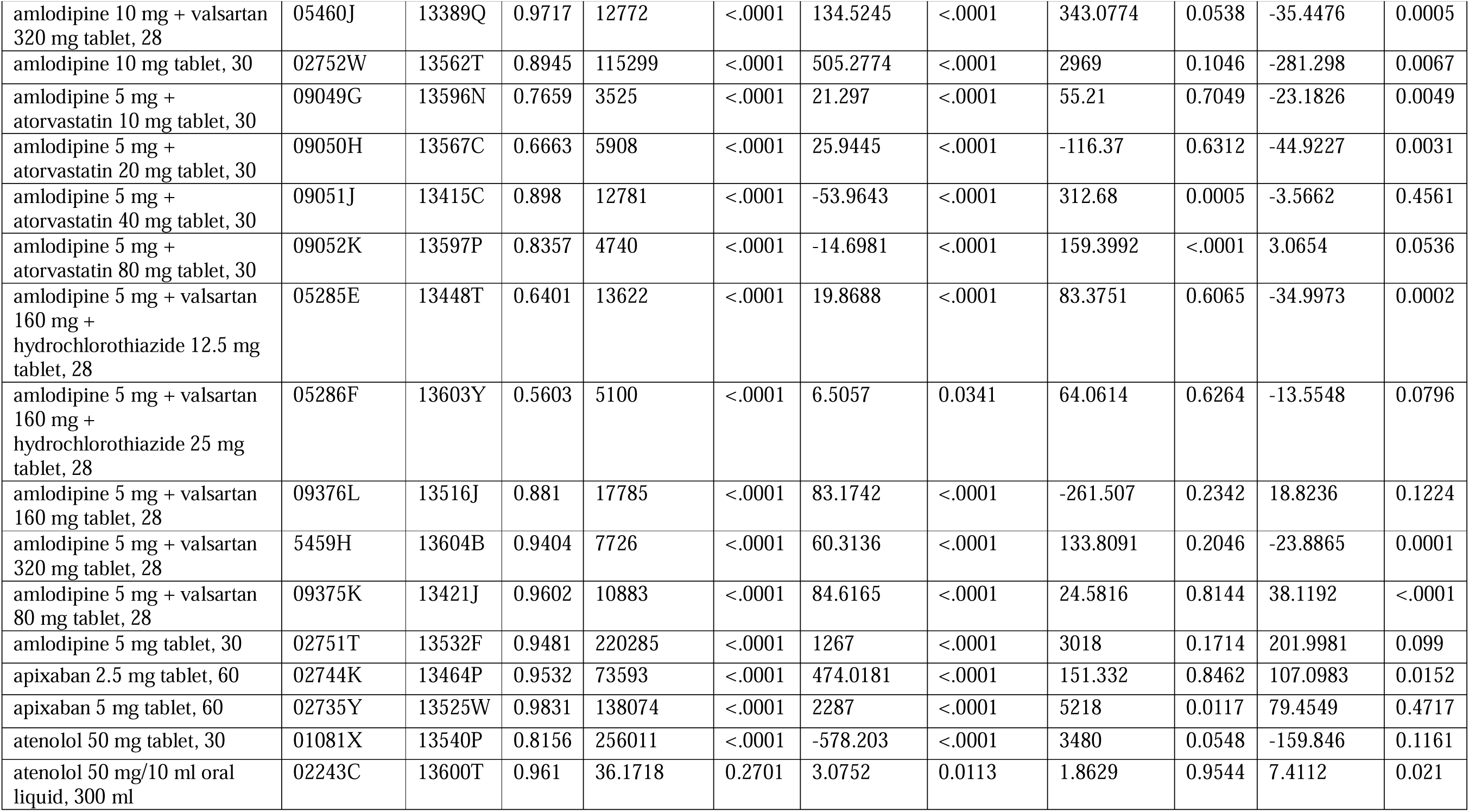
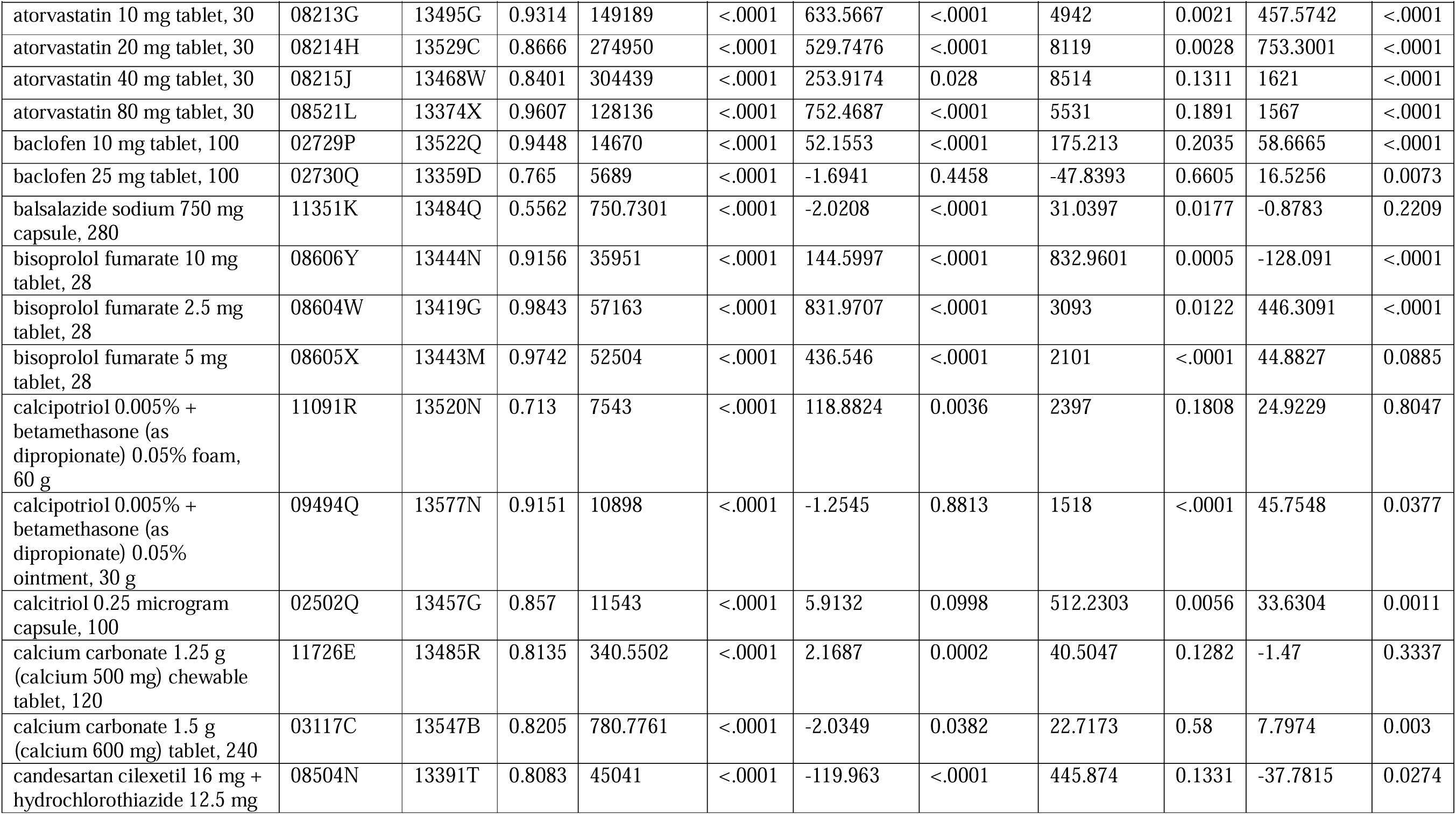
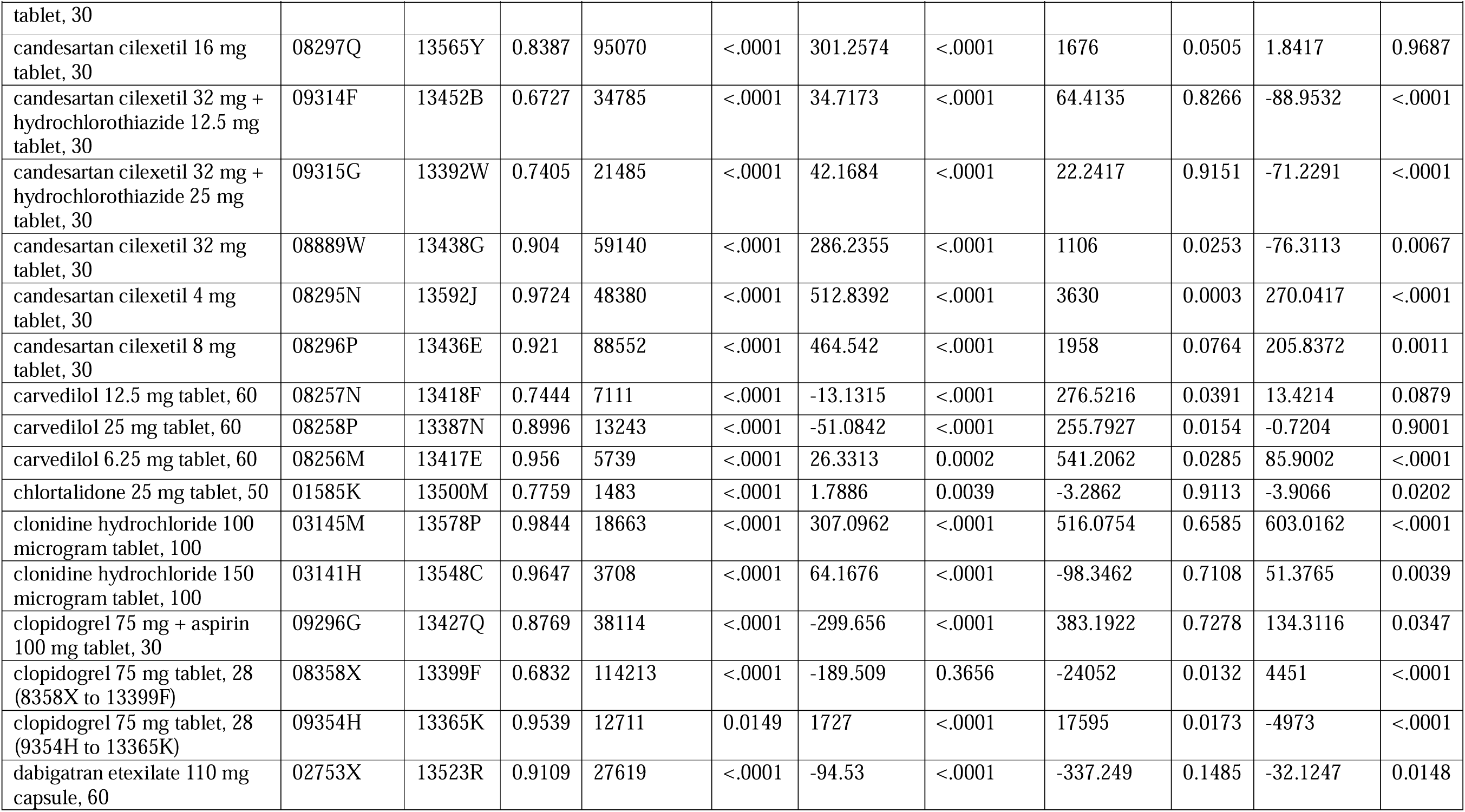
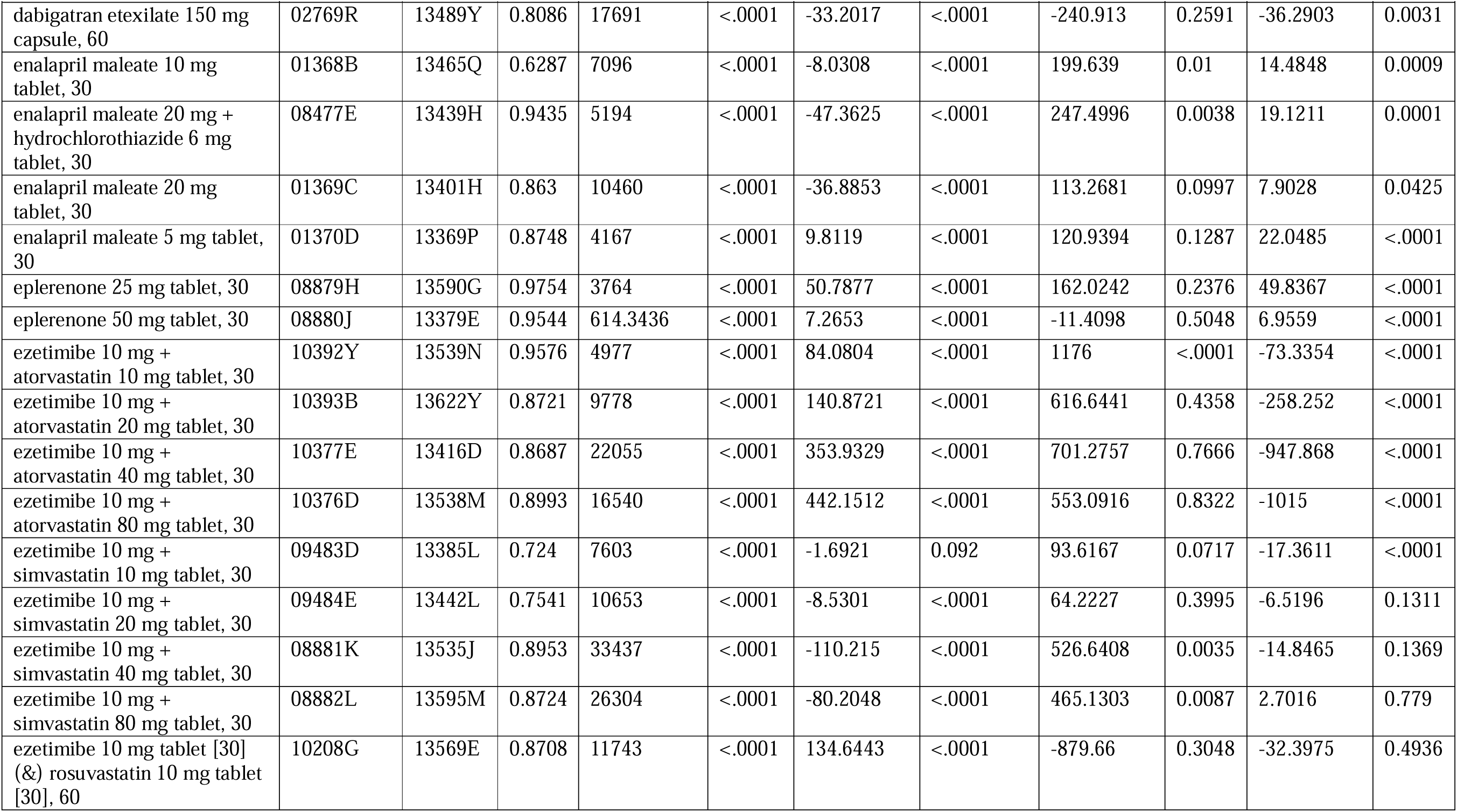
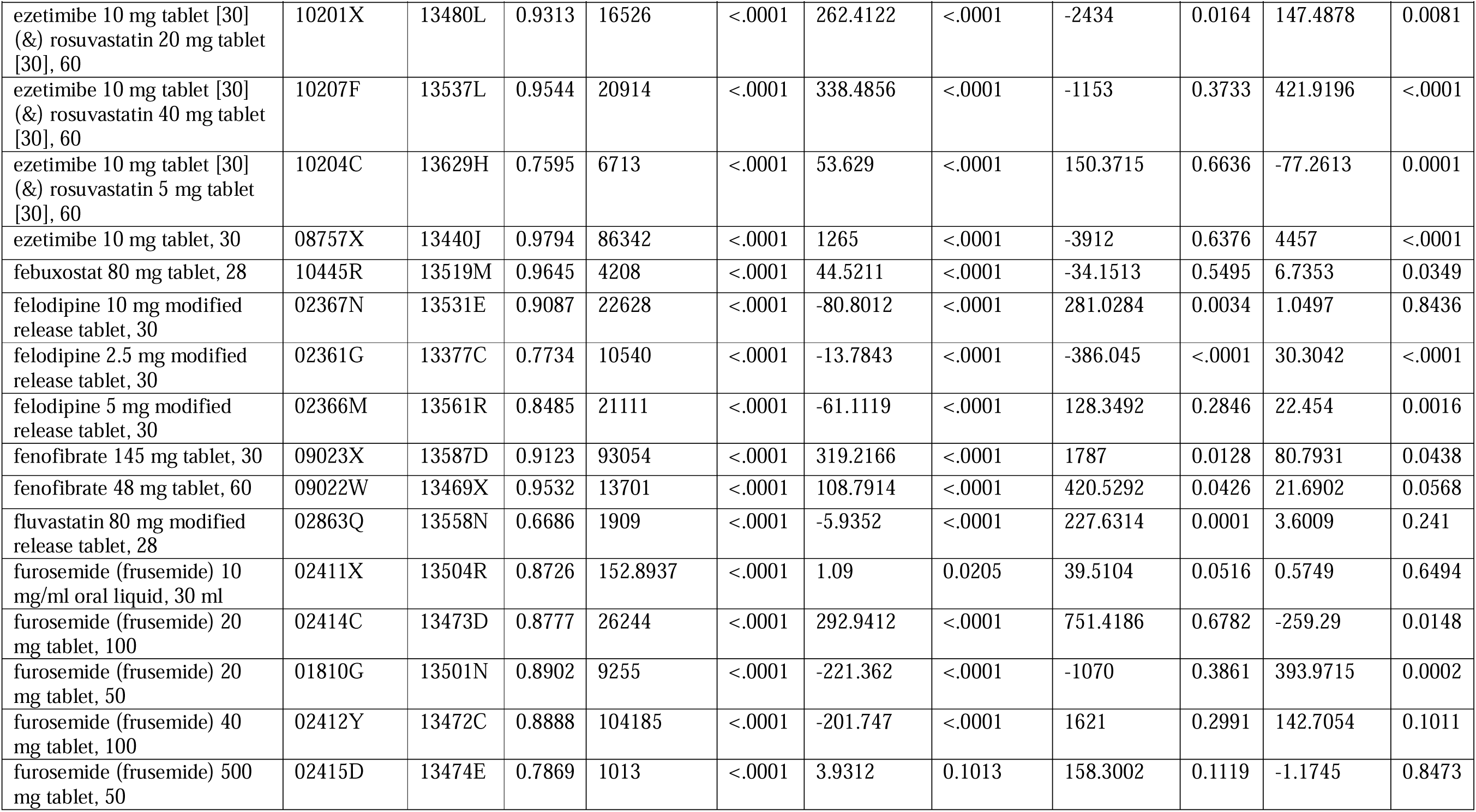
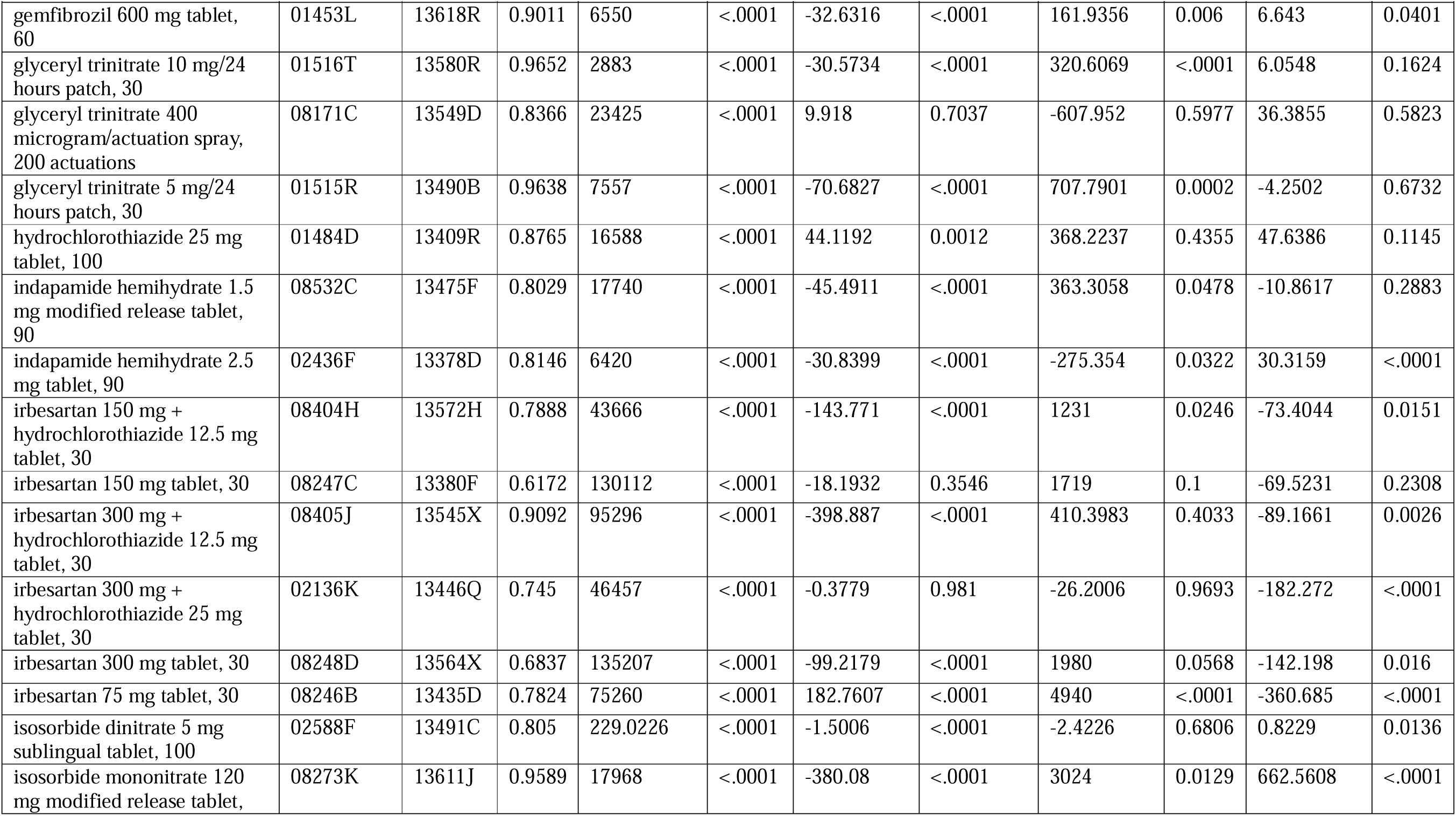
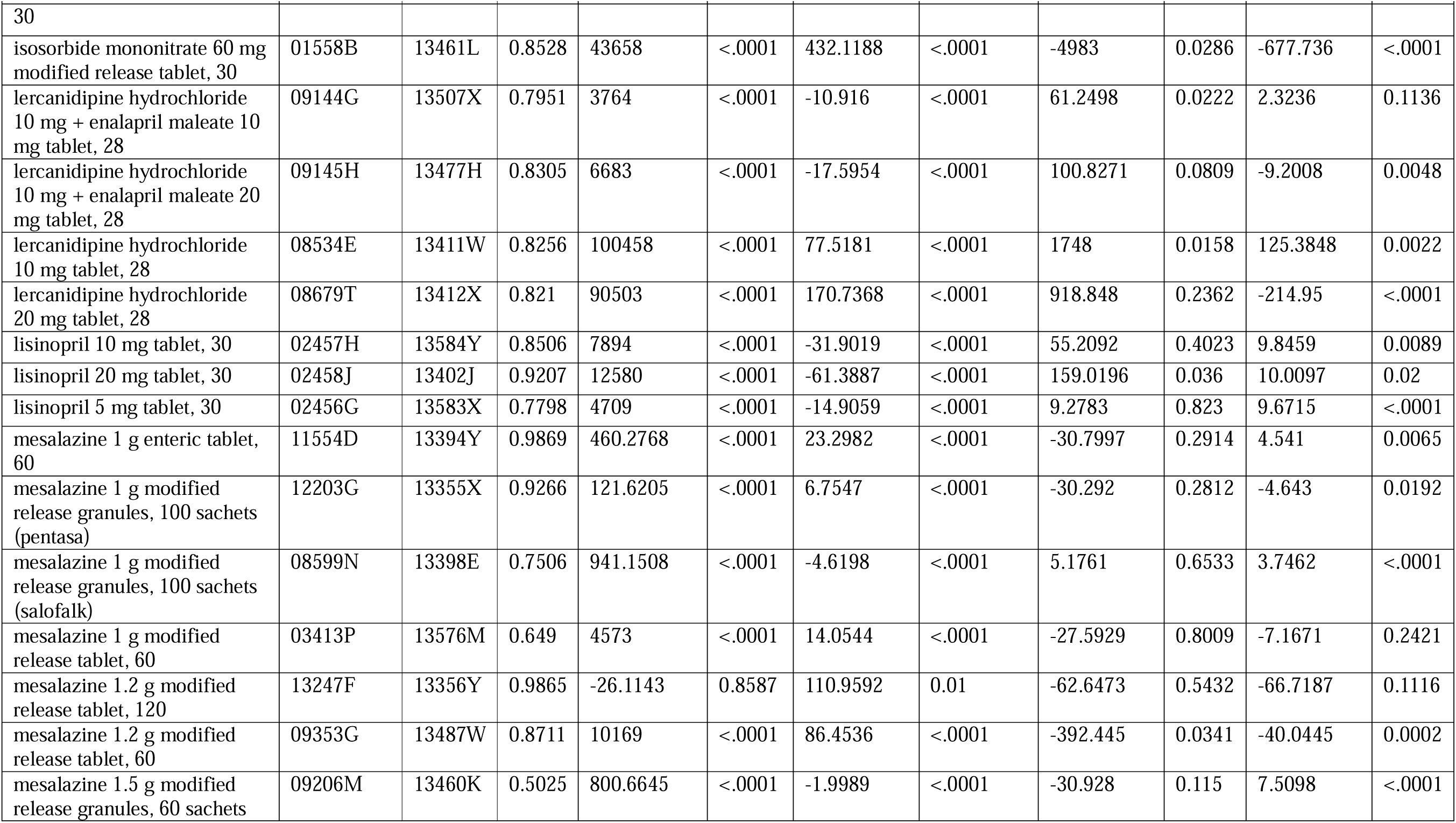
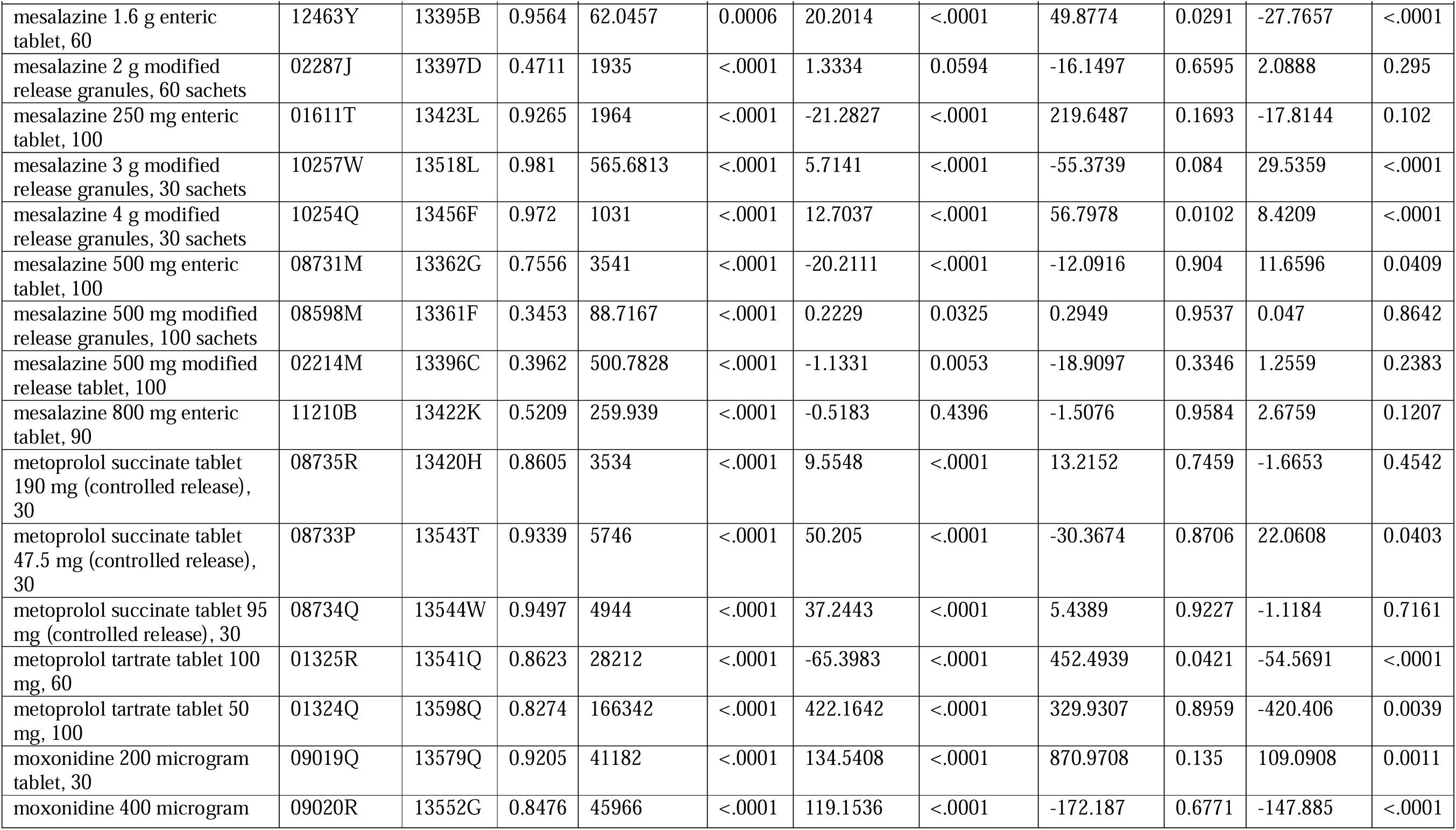
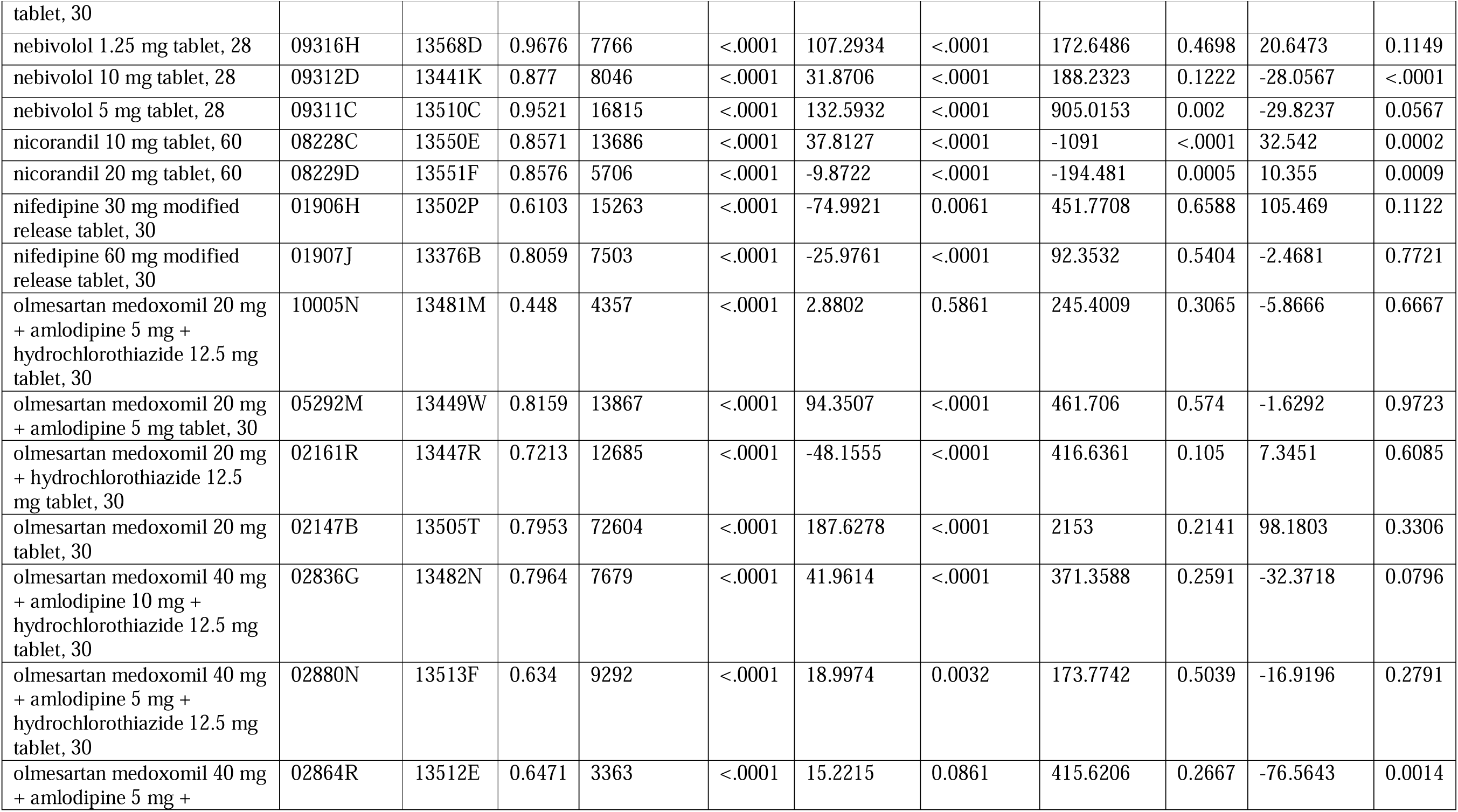
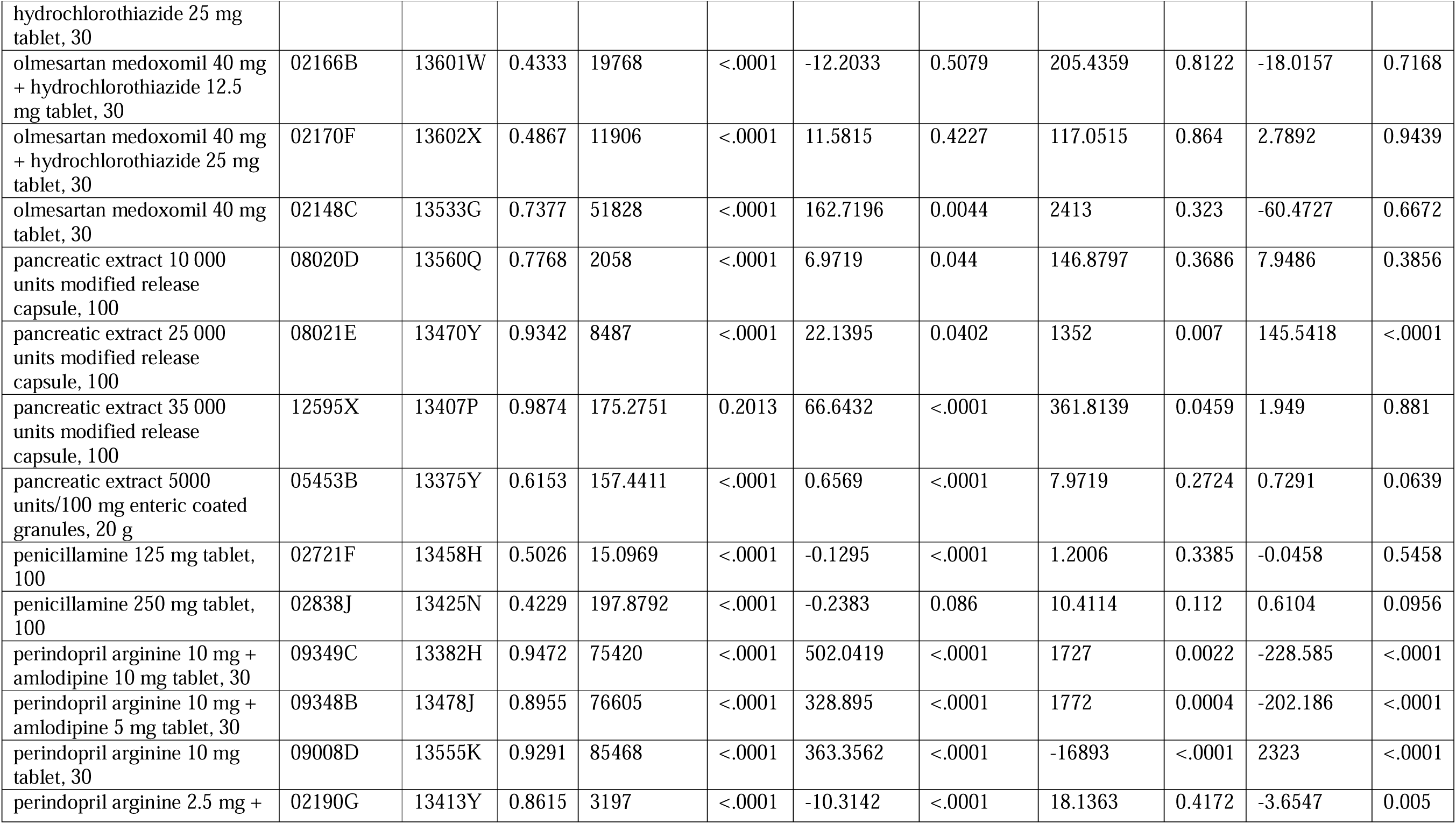
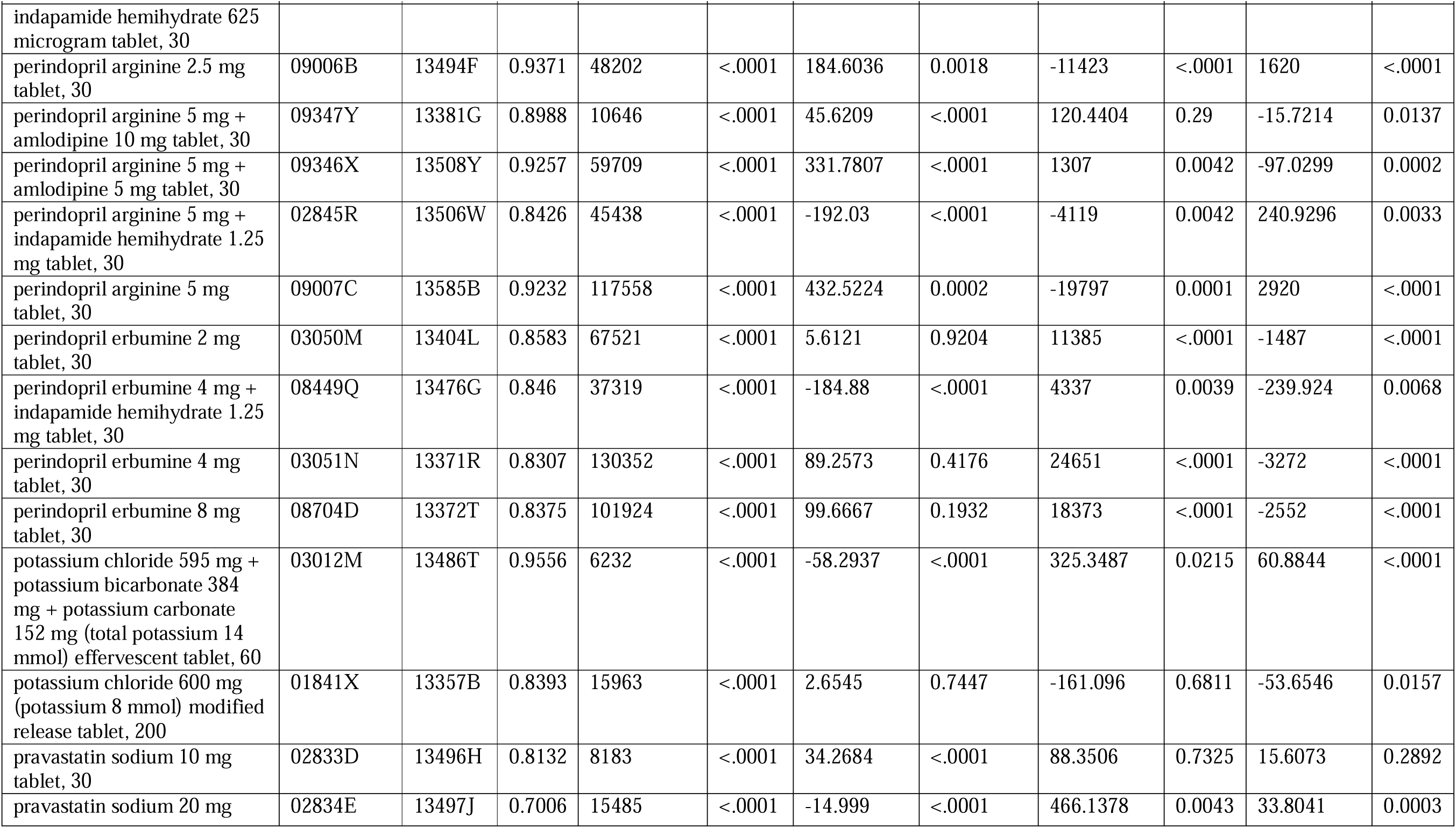
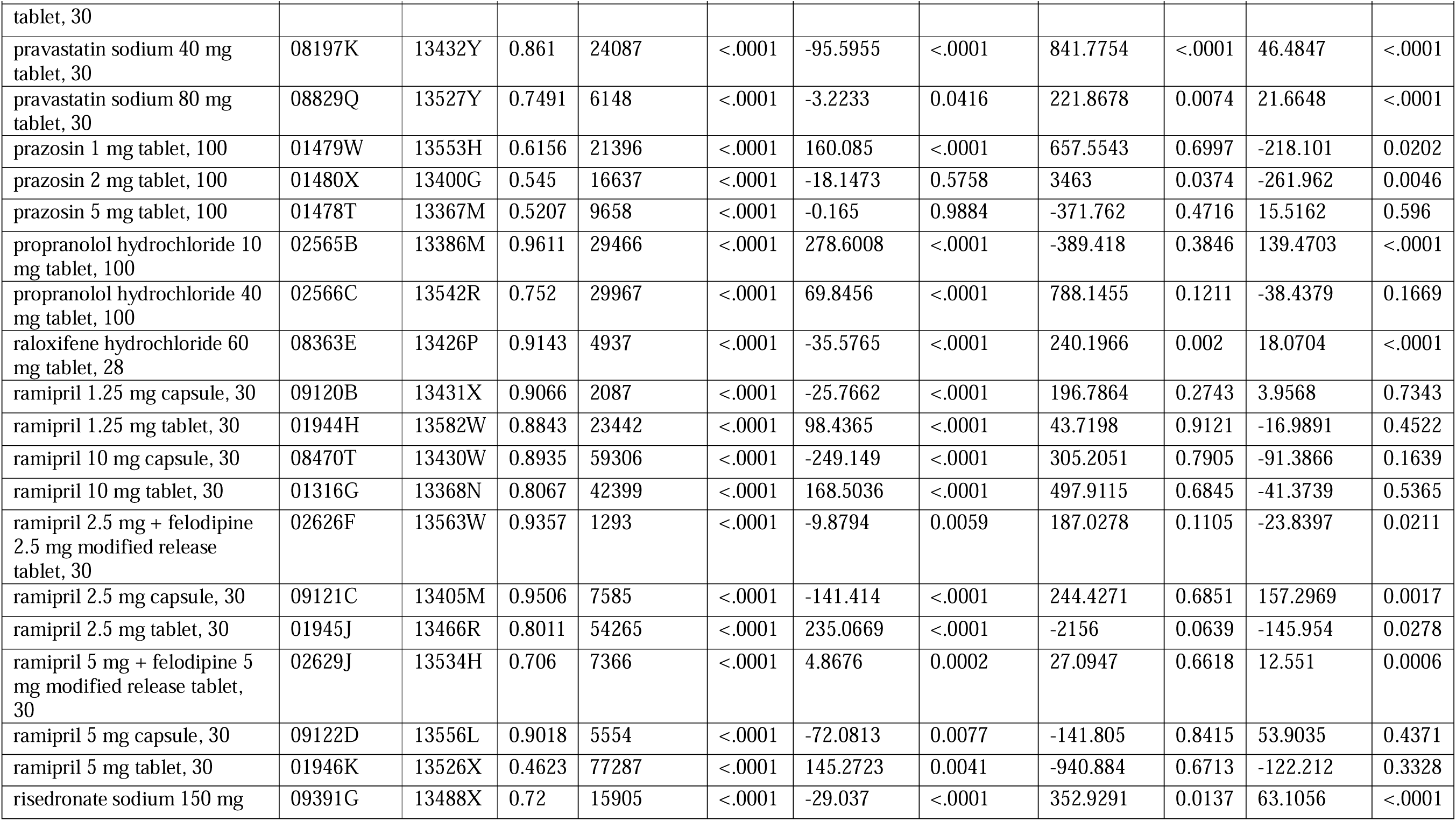
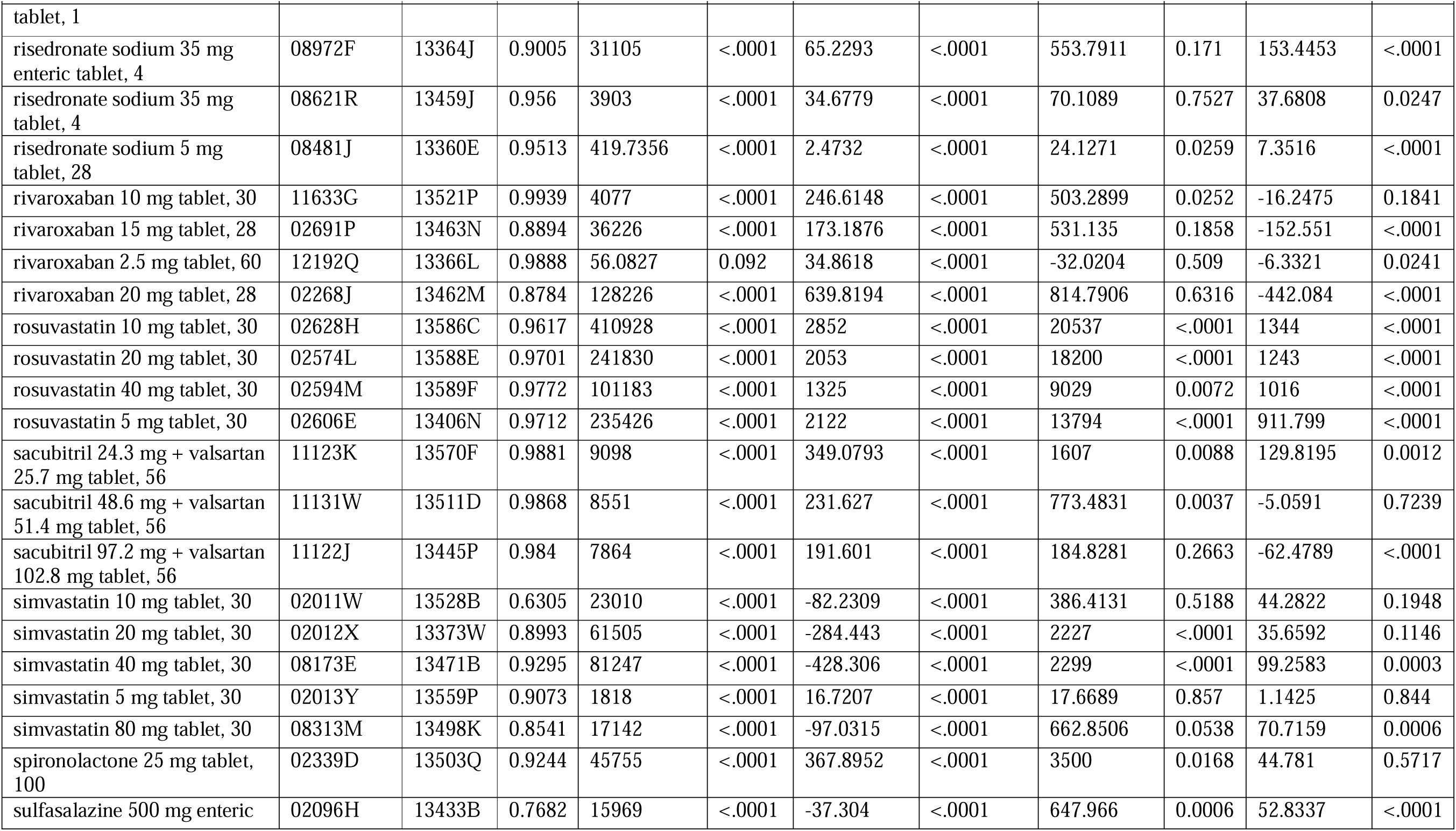
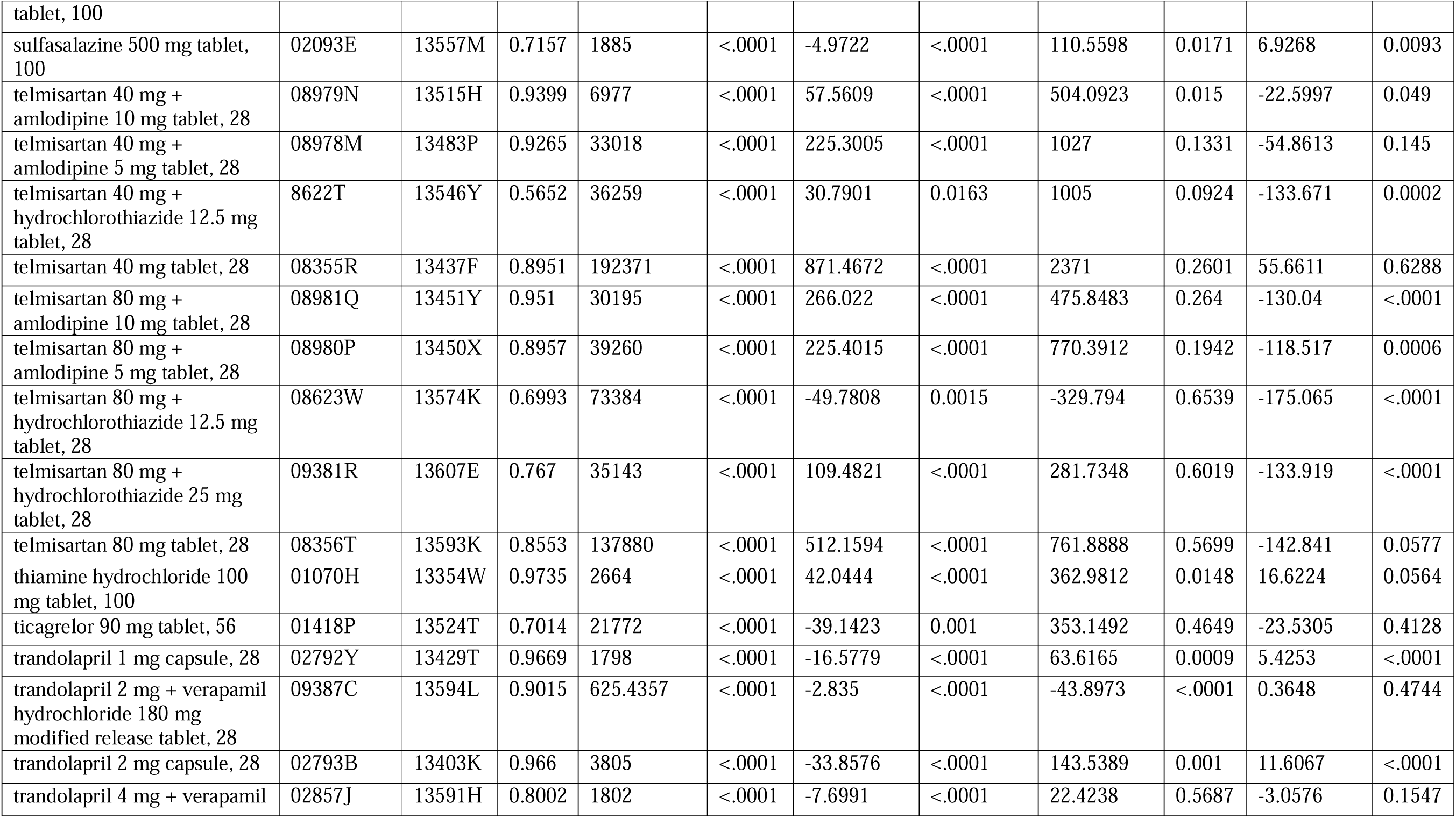
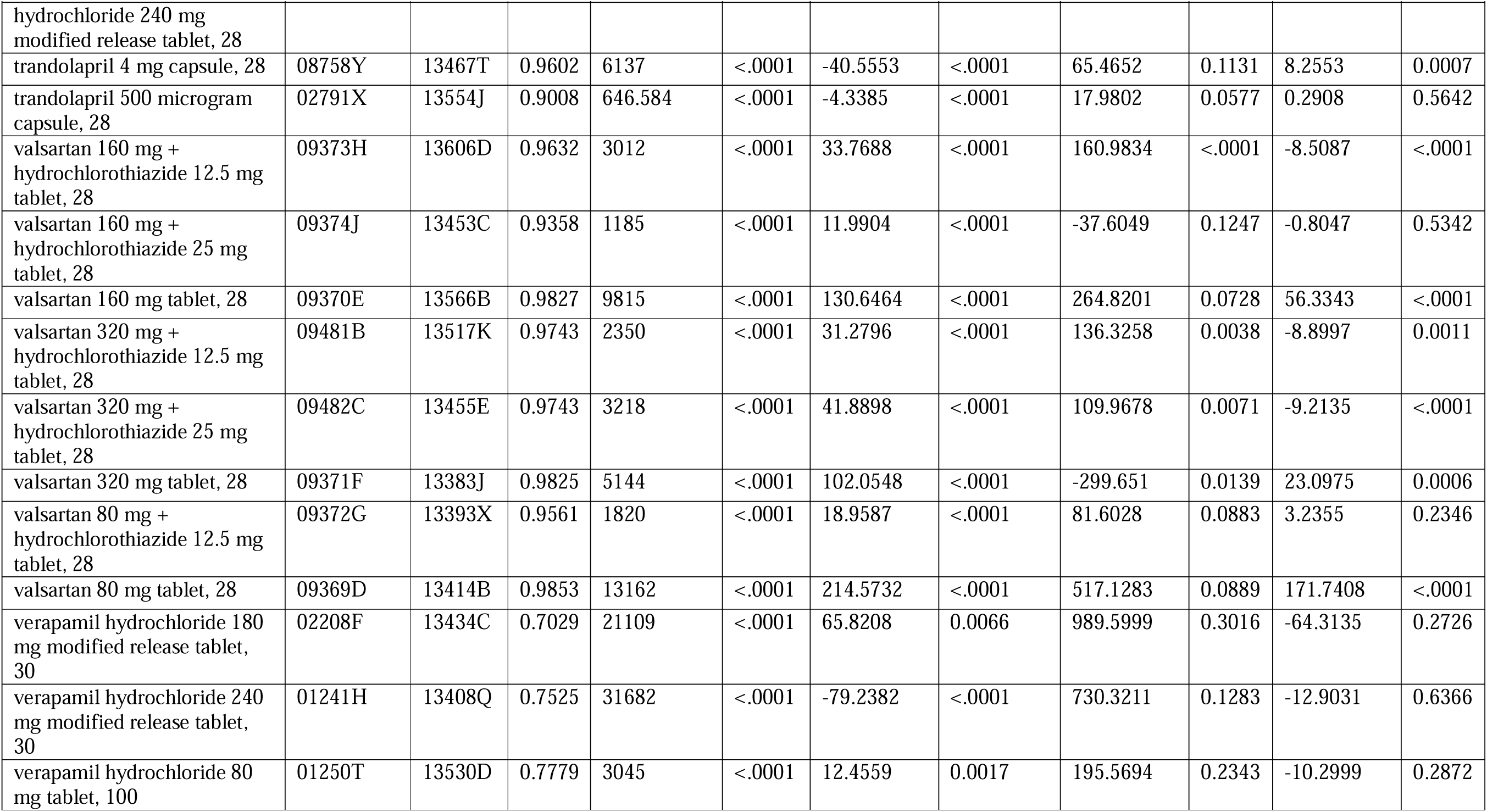
Interrupted time series segmented regression outputs for each Stage 1 medicine.

**Appendix Table 6.**
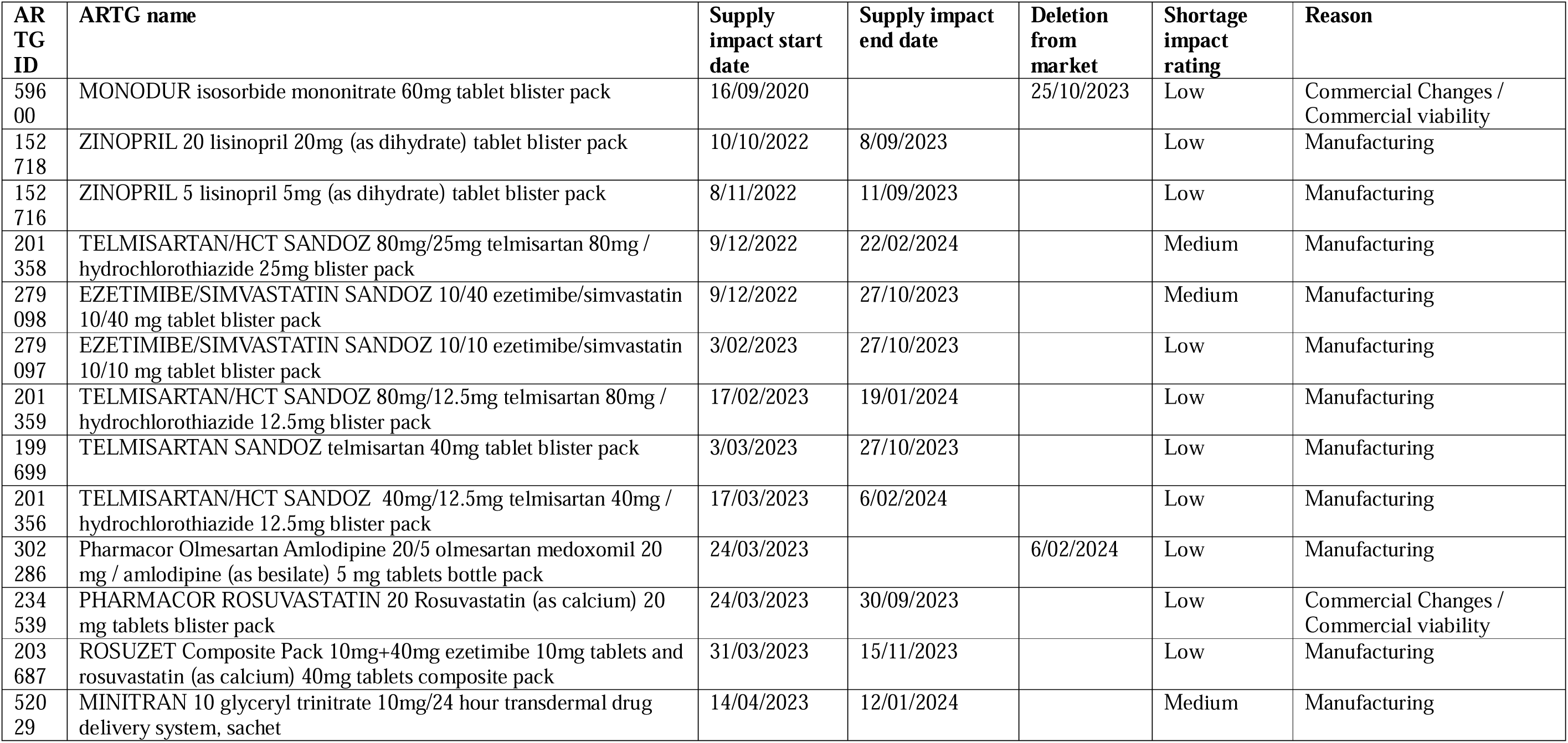
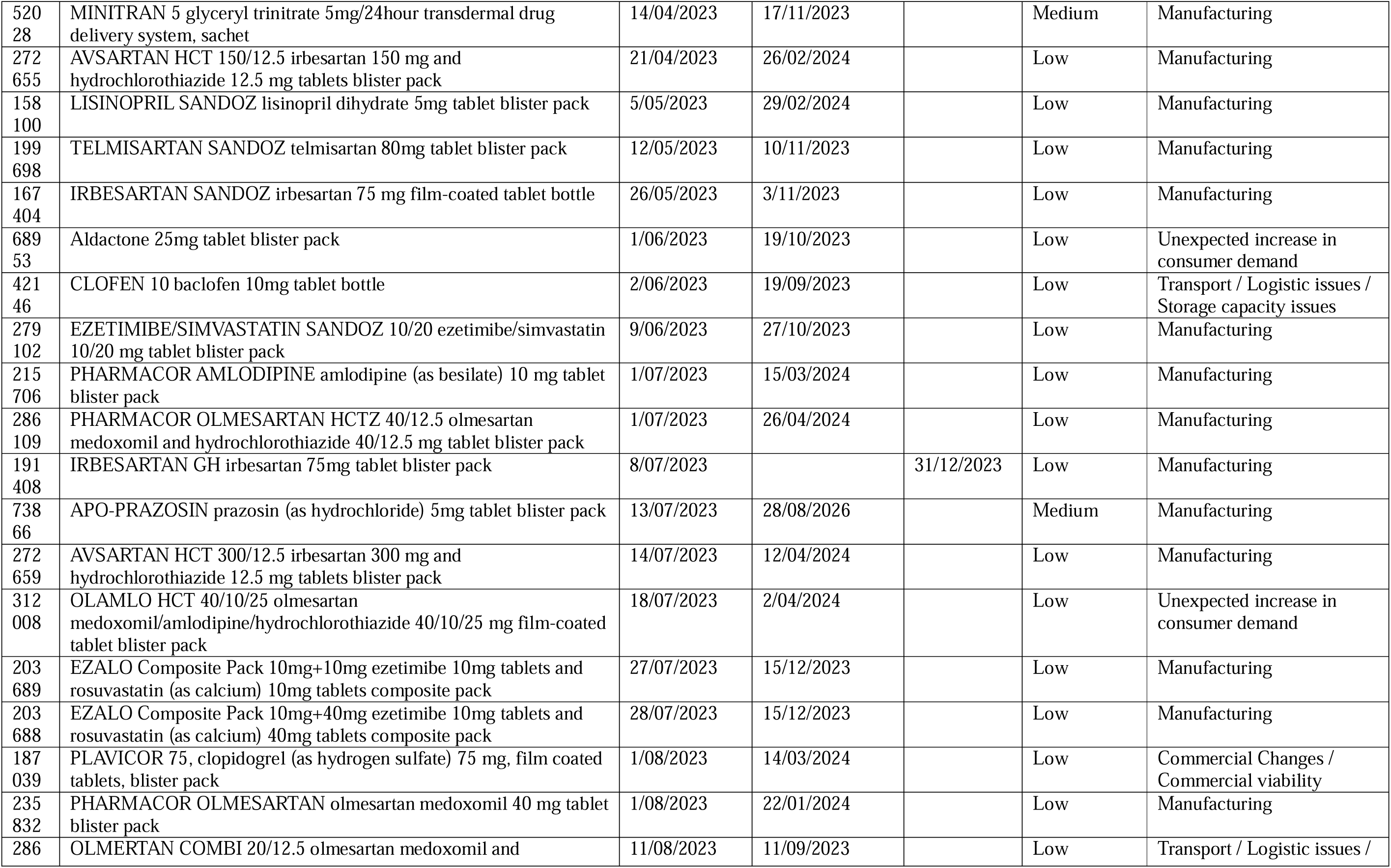
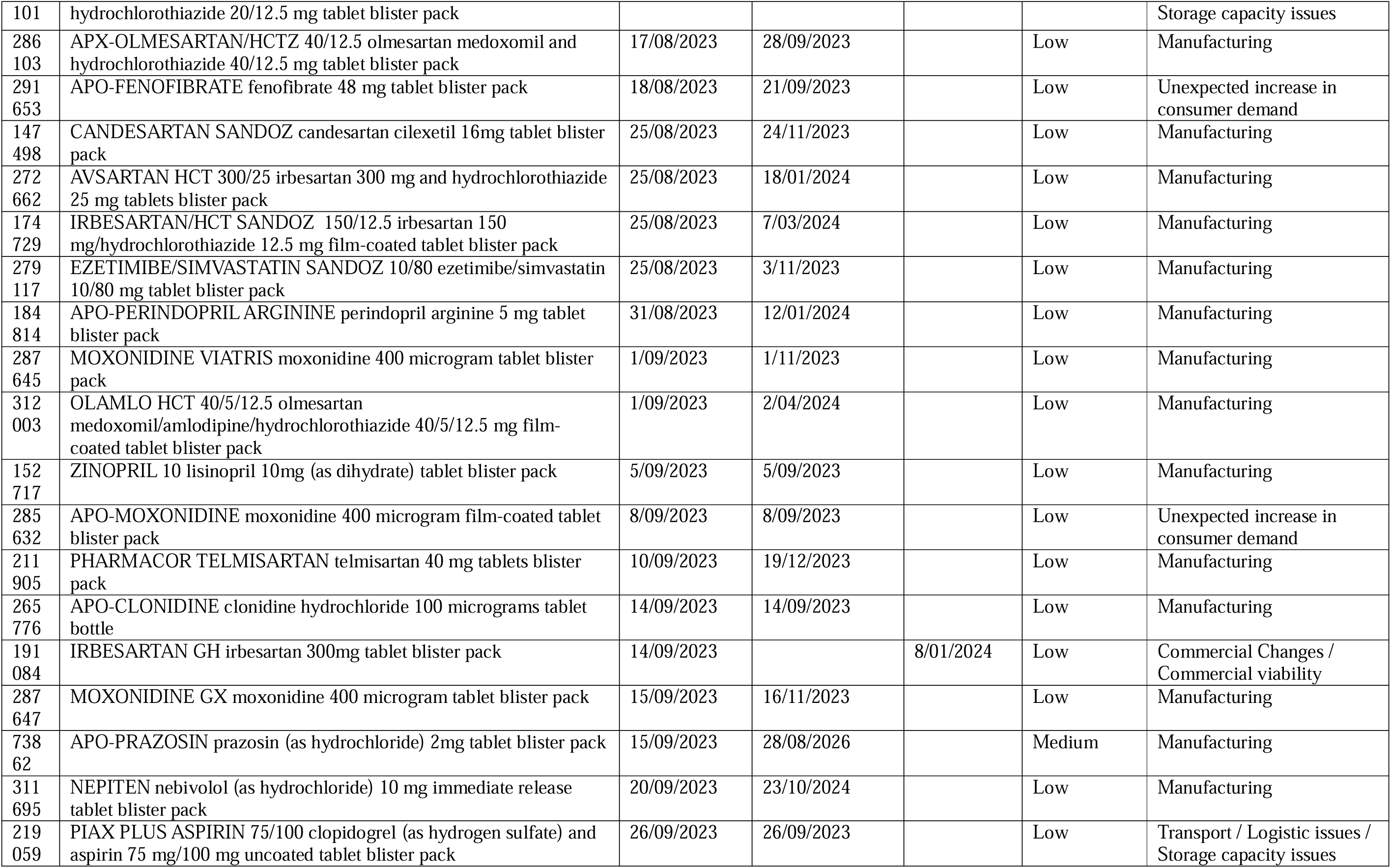
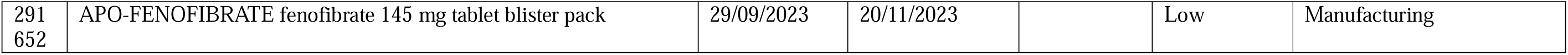
Medicine shortages of 60-day medicines from the Therapeutic Goods Administration Medicines shortages database. This table includes shortage data where the medicine was in shortage in September 2023.

